# A regenerative stem cell phenotype marked by ANXA1 defines aggressive colorectal cancer biology

**DOI:** 10.64898/2026.10.28.26366700

**Authors:** Henry Rotte, Enric Domingo, Gabriel Valbuena, Nadia Nasreddin, Andrea Harkin, Tim Iveson, Mark P Saunders, Karin A Oien, Noori Maka, Francesco Pezzella, Leticia Campo, Molly Browne, Mark Glaire, Jennifer Hay, Joanne Edwards, Owen J Sansom, Caroline Kelly, Ian Tomlinson, Rachel Kerr, David Kerr, TransSCOT consortium, imCMS Consortium, Simon Leedham, David N Church, Konstantin Bräutigam, Viktor H Koelzer

**Author notes:** HR and ED are joint first authors. KB and VHK are joint last authors.

## Abstract

**Background:** LGR5 and ANXA1 are markers for specific intestinal stem cell states. LGR5^+^/ANXA1^−^ crypt base columnar cells (CBCs) maintain homeostasis, whereas ANXA1^+^/LGR5^−^ regenerative stem cells (RSCs) are present in fetal and repairing epithelium. Both states have been described in colorectal cancer (CRC), yet their clinical significance remains unclear. We aimed to determine how ANXA1 and LGR5 expression relates to molecular subtype, immune microenvironment and outcome in CRC.

**Methods:** Dual-colour in situ hybridisation for ANXA1 and LGR5 was performed on tissue microarrays from 2163 patients in the randomised SCOT trial (ISRCTN59757862). AI-based digital pathology quantified marker-positive area by compartment and image based consensus molecular subtype (imCMS). Markers were related to clinicopathological, molecular and immune data, and to survival using Kaplan-Meier and multivariable Cox analyses.

**Results:** ANXA1 and LGR5 showed inverse enrichment among a diverse group of parameters. ANXA1 was highest in CMS1 and LGR5 in CMS2, with CMS3 and CMS4 being intermediate. ANXA1 dominance was associated with right-sidedness, mismatch repair deficiency, BRAF-V600E mutation, high stromal content, T4 stage, low stromal FoxP3 and high epithelial CD68 density. Combining both markers improved prognostic discrimination compared with either alone (cancer-specific survival p<0.001; recurrence-free interval p=0.004), with ANXA1 dominance remaining an independent predictor of recurrence after adjustment (HR 1.43, 95% CI 1.08-1.90, p=0.014).

**Conclusion:** Relative ANXA1 enrichment defines an aggressive, stroma-rich, macrophage-enriched CRC subgroup with poor prognosis.

## Introduction

Colorectal cancer (CRC) is the cause of significant morbidity and mortality. A better understanding of the pathophysiology of CRC has led to new therapeutic options, such as immunotherapy (1). The role of the tumour immune microenvironment (TIME) and cancer stem cells (CSCs) is increasingly becoming the focus of research (2,3).

A major advance in understanding the molecular basis of CRC was the introduction of the Consensus Molecular Subtypes (CMS), which categorize CRC into four molecular subtypes, CMS1 to CMS4, each characterized by specific molecular features (4). CMS1 is understood to be “immune-hot” with MMR deficiency, BRAF-V600E mutations and response to PD1/PD-L1-inhibitors, with more favourable prognosis in early disease and poor prognosis in advanced stages (4). CMS2 is labeled “canonical”, and is characterised by chromosomal instability, microsatellite stability, left-sided predominance and activation of the WNT and MYC pathways. CMS3 is characterised by prominent metabolic dysregulation, frequent KRAS mutations, high molecular heterogeneity and intermediate prognosis. Lastly, CMS4 is characterised by stromal invasion and TGF-β signalling and has the poorest prognosis. More recently, a refined CMS classification has been proposed, focusing on epithelial cell diversity and reducing the CMS into two epithelial subtypes, labelled iCMS2 and iCMS3 (5). CMS1 and CMS3 cancers map almost exclusively to the iCMS3 category. CMS2 maps onto the iCMS2 category, whereas CMS4 tumours are roughly split between iCMS2 and iCMS3 epithelium, indicating that the stromal features defining CMS4 are independent of the underlying epithelial programme.

Beyond molecular classification, growing attention has focused on interactions between cancer and the immune system, with immune evasion recognized as a “hallmark of cancer” (6). Previous investigations into the TIME in CRC have established high intraepithelial CD8^+^ and high intrastromal FoxP3^+^ cell counts as protective (7–9). The favourable effect of FoxP3^+^ T-regulatory cells in CRC is paradoxical in comparison to most other carcinomas and probably reflects specific FoxP3^+^ subpopulations (10). Macrophages show a more nuanced picture. M2-like macrophages generally favour tumour growth, whereas M1-like macrophages are more tumour-suppressive (11). Research also indicates that CD20^+^ B-cells are a favourable prognostic marker in CRC (12).

Cancer stem cells (CSCs) play an important role in CRC development and progression (13). Instead of distinct categories, CSCs represent a phenotypic spectrum, and can adapt to environmental pressure. Two stem cell modes were proposed on this spectrum to represent an optimum fitness landscape for cancer. These were labelled Regenerative Stem Cells (RSC) and Crypt Base Columnar (CBC) stem cells. CBCs are identified by *Leucine-rich repeat-containing G-protein coupled receptor 5* (LGR5) positivity (14,15), and have been demonstrated to be responsible for steady-state homeostasis in mature intestinal tissue (16). Using ISH in a smaller cohort, LGR5 expression has been described in both colorectal adenomas and carcinomas (17) with observed LGR5 expansion in CRC (18). On the contrary, RSCs are defined by the activation of YAP/TAZ signaling, as well as fetal gene expression (19), and can be identified in part through *Annexin A1* (ANXA1). Stem cells with an RSC signature are thought to have a physiological role in tissue regeneration after injury and ANXA1 has been shown to be upregulated in fetal and repairing epithelium (20). ANXA1 and LGR5 are inversely proportionally enriched, therefore representing distinct adaptive strategies of CRC (15). While LGR5 is a well established stem cell marker (21), the role of ANXA1 is more complex. ANXA1 was first investigated for its role as an effector of glucocorticoid signaling and in inflammation-resolution (22,23). Research on ANXA1 in cancer has shown it to have a context and localisation dependent role, with generally antiproliferative effects in early stage disease, while facilitating metastasis in later disease (24).

To map the spatial expression of the stem cell markers ANXA1 and LGR5 in CRC comprehensively, we used tissue microarrays from the SCOT trial cohort; this large randomised controlled study compared 3 months of oxaliplatin-based chemotherapy to the traditional 6 months for most high-risk stage II and stage III CRC patients and involved 244 academic centres and 6,088 CRC patients (25,26). We hypothesised that ANXA1 and LGR5 expression, reflecting the admixture of RSC and CBC stem cells, would correlate with the clinical, pathological, and molecular features of CRC and influence survival outcomes.

## Materials and Methods

### SCOT trial cohort

This study is a prospective–retrospective correlative analysis of tumour tissue from SCOT (ISRCTN59757862), an international, randomised, phase III non-inferiority trial comparing 3 versus 6 months of adjuvant oxaliplatin-based chemotherapy in patients with resected stage III or high-risk stage II colorectal cancer (25,26). High-risk stage II disease was defined as one or more of: T4 disease, tumour obstruction with or without perforation, fewer than 10 lymph nodes harvested, poorly differentiated histology, perineural invasion, or extramural venous or lymphatic vascular invasion. Between March 27, 2008, and November 29, 2013, 6,088 patients were randomised to 3 or 6 months of treatment, with allocation by minimization. The regimen (CAPOX or FOLFOX) was selected before randomisation by the patient and treating physician. tumour tissue was collected prospectively at participating centers within the TransSCOT translational substudy.

Of the 6,088 patients enrolled, 2163 patients had tissue samples available for analysis. (**Figure 1**). Of the 6088 Patients, 3798 had no tissue samples available for evaluation and a further 123 patients had no corresponding imCMS data available. 9 Cores were found to contain no usable tissue (stroma or epithelium), but instead only erythrocytes, necrosis and/or artifacts and were therefore also excluded. The sample size was determined by tissue availability; no formal power calculation was performed. Both sexes were included in analysis and sex was considered as a covariate in all multivariable models. Clinicopathological, molecular and immune variables are shown in (**Tables 1-3**). Mean follow-up for overall survival (OS), cancer-specific survival (CSS), recurrence-free interval (RFI) and survival after relapse (SAR) was 5.58, 5.63, 5.01 and 2.36 years respectively. Consensus molecular subtyping was performed by imCMS v1.5 as described in the digital pathology section.

**Figure 1.**
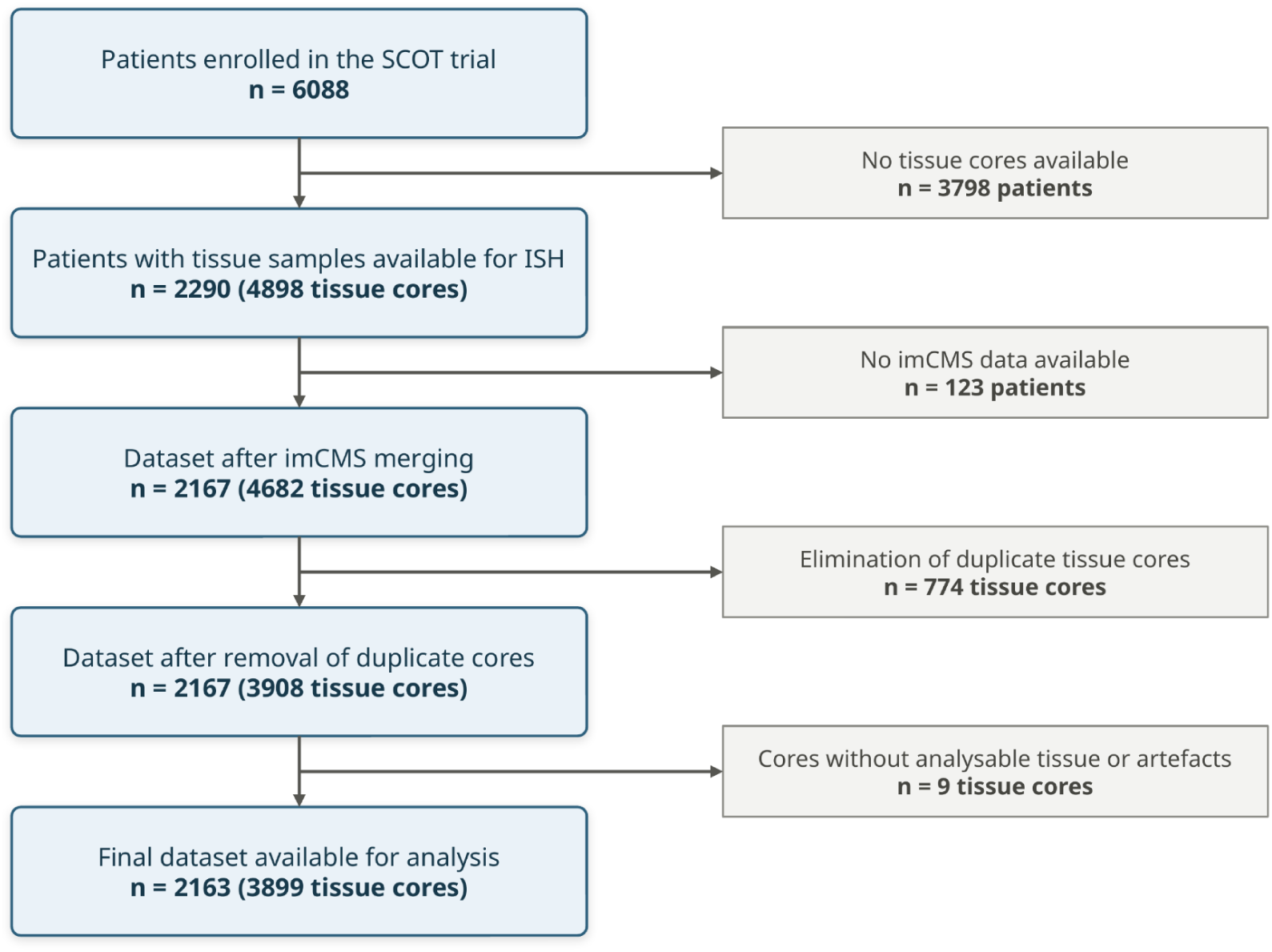
Flow (CONSORT) diagram of SCOT trial cases included in this study.

**Table 1.** ANXA1 expression in the tumour centre (TC) and clinical parameters. ANXA1, Annexin A1; CAPOX, capecitabine+oxaliplatin; CD, cluster of differentiation; FOLFOX, Folinic Acid+Fluorouracil (5-FU)+Oxaliplatin; IM, invasive margin; imCMS, image based consensus molecular subtype; LGR5, leucine-rich repeat-containing G-protein coupled receptor 5; MMR, mismatch repair; N, number (sample size); Q, quartile; RFI, recurrence-free interval; TC, tumour centre; Tumour proportion, equals 1-TSR

| Characteristic | Q1<br>N = 503<sup>a</sup> | Q2<br>N = 503<sup>a</sup> | Q3<br>N = 502<sup>a</sup> | Q4<br>N = 503<sup>a</sup> | p value<sup>b</sup> |
| --- | --- | --- | --- | --- | --- |
| Age | 65 (58, 70) | 65 (58, 70) | 65 (58, 70) | 64 (59, 70) | >0.9 |
| **Sex** |  |  |  |  | 0.086 |
| Female | 189 (23%) | 188 (23%) | 210 (26%) | 221 (27%) |  |
| Male | 314 (26%) | 315 (26%) | 292 (24%) | 282 (23%) |  |
| Performance status | 148 (25%) | 156 (26%) | 131 (22%) | 161 (27%) | 0.2 |
| **Sidedness** |  |  |  |  | <0.001 |
| Left | 312 (27%) | 311 (26%) | 299 (25%) | 255 (22%) |  |
| Right | 189 (23%) | 184 (23%) | 197 (24%) | 241 (30%) |  |
| **Trial arm** |  |  |  |  | 0.3 |
| 12 weeks | 242 (24%) | 250 (25%) | 244 (24%) | 269 (27%) |  |
| 24 weeks | 261 (26%) | 253 (25%) | 258 (26%) | 234 (23%) |  |
| **Planned treatment** |  |  |  |  | 0.6 |
| CAPOX | 363 (26%) | 362 (25%) | 352 (25%) | 346 (24%) |  |
| FOLFOX | 140 (24%) | 141 (24%) | 150 (26%) | 157 (27%) |  |
| T4 stage | 160 (24%) | 154 (23%) | 174 (26%) | 184 (27%) | 0.2 |
| **Nodal stage** |  |  |  |  | 0.3 |
| 0 | 112 (28%) | 98 (24%) | 94 (23%) | 103 (25%) |  |
| 1 | 284 (26%) | 278 (25%) | 274 (25%) | 264 (24%) |  |
| 2 | 107 (21%) | 127 (25%) | 134 (27%) | 136 (27%) |  |
| Tumour proportion | 84 (66, 96) | 69 (48, 84) | 58 (38, 77) | 51 (27, 73) | <0.001 |
| CD8, intraepithelial (z-score) | 0.11 (-0.43, 0.62) | 0.19 (-0.47, 0.68) | 0.16 (-0.35, 0.65) | 0.20 (-0.42, 0.71) | 0.6 |
| CD8, intrastromal (z-score) | 0.12 (-0.50, 0.61) | 0.08 (-0.57, 0.62) | 0.11 (-0.52, 0.65) | 0.15 (-0.41, 0.70) | 0.5 |
| FOXP3, intraepithelial (z-score) | 0.17 (-0.35, 0.54) | 0.20 (-0.21, 0.59) | 0.19 (-0.26, 0.58) | 0.22 (-0.33, 0.60) | 0.4 |
| FOXP3, intrastromal (z-score) | 0.17 (-0.63, 0.68) | 0.22 (-0.40, 0.71) | 0.26 (-0.42, 0.80) | 0.08 (-0.65, 0.65) | 0.004 |
| CD20, intraepithelial (z-score) | 0.25 (-0.41, 0.70) | 0.21 (-0.49, 0.71) | 0.22 (-0.52, 0.74) | 0.11 (-0.81, 0.68) | 0.2 |
| CD20, intrastromal (z-score) | 0.06 (-0.50, 0.57) | 0.14 (-0.44, 0.63) | 0.19 (-0.41, 0.66) | 0.12 (-0.45, 0.59) | 0.3 |
| CD68, intraepithelial (z-score) | -0.11 (-0.68, 0.61) | 0.01 (-0.53, 0.64) | 0.03 (-0.53, 0.65) | 0.23 (-0.35, 0.86) | <0.001 |
| CD68, intrastromal (z-score) | 0.05 (-0.44, 0.46) | 0.09 (-0.35, 0.52) | 0.11 (-0.30, 0.50) | 0.14 (-0.36, 0.56) | 0.15 |
| **FOXP3 (intrastromal) / CD8 (intraepithelial)** |  |  |  |  | 0.8 |
| high/high | 150 (24%) | 166 (26%) | 170 (27%) | 152 (24%) |  |
| high/low | 191 (26%) | 182 (25%) | 181 (25%) | 184 (25%) |  |
| low/low | 161 (26%) | 154 (24%) | 150 (24%) | 165 (26%) |  |
| **imCMS** |  |  |  |  | <0.001 |
| imCMS1 | 84 (17%) | 97 (20%) | 101 (21%) | 206 (42%) |  |
| imCMS2 | 253 (35%) | 200 (28%) | 173 (24%) | 99 (14%) |  |
| imCMS3 | 55 (20%) | 77 (28%) | 75 (28%) | 65 (24%) |  |
| imCMS4 | 99 (21%) | 114 (24%) | 136 (29%) | 117 (25%) |  |
| Mixed | 12 (20%) | 15 (25%) | 17 (28%) | 16 (27%) |  |
| **MMR status** |  |  |  |  | <0.001 |
| Deficient | 44 (21%) | 37 (18%) | 43 (21%) | 84 (40%) |  |
| Proficient | 393 (26%) | 409 (27%) | 388 (25%) | 348 (23%) |  |
| **BRAF V600E** |  |  |  |  | 0.080 |
| Mutant | 8 (16%) | 11 (22%) | 10 (20%) | 21 (42%) |  |
| Wild-type | 100 (27%) | 91 (25%) | 84 (23%) | 94 (25%) |  |
| **KRAS** |  |  |  |  | 0.4 |
| Mutant | 40 (29%) | 31 (22%) | 26 (19%) | 42 (30%) |  |
| Wild-type | 68 (24%) | 71 (25%) | 68 (24%) | 73 (26%) |  |

<sup>a</sup>Median (Q1, Q3); n (%).
<sup>b</sup>Kruskal–Wallis rank sum test; Pearson’s chi-squared test.

For the clinical endpoints, time was measured from randomisation, or from trial registration for patients randomised after 3 months of therapy. Overall survival (OS) was defined as the time to death from any cause. Cancer-specific survival (CSS) was defined as the time to death from colorectal cancer, with censoring at last contact or at death from another cause. Recurrence-free interval (RFI) was defined as the time to relapse, with censoring at last contact or at death without recurrence. Survival after relapse (SAR) was defined as the time from first relapse to death from any cause in patients who relapsed, with censoring at last contact.

### Tissue microarrays (TMAs)

Twenty-five pathologist-annotated TMAs were analyzed as previously described (8). TMAs were constructed from 0.6-mm cores punched from formalin-fixed paraffin-embedded (FFPE) primary tumour blocks and stored at room temperature, protected from air (8). Cores were sampled from the tumour center (TC) and invasive margin (IM). The final dataset contained 3899 tissue cores. Tissue content per core was calculated by *(stroma + tumour epithelium) / total classified area* and varied between 0% and 70.7%. The rest of the area was composed of white space, necrosis, erythrocytes or artefacts. Average analysable tissue per core was 32.3% of classified area, with 95% of tissue cores having between 4.1% and 52.1% analysable tissue. The total classified area per tissue core varied between 0.681mm² and 0.960mm².

### In-situ hybridisation (ISH) and immune phenotyping

Multiplex in situ hybridisation (ISH) of ANXA1 (human; RNAscope™ Probe - Hs-ANXA1; yellow chromogen, Catalog # 465411) and LGR5 (human; RNAscope™ Probe - LGR5, Catalog # 311021-C2; red chromogen) was performed using the RNAscope™ Multiplex Fluorescent Assay, following the manufacturer’s instructions (Bio-Techne, Minnesota, United States). Sections were baked at 60 °C for 1 h, then dewaxed in xylene and ethanol. Multiplex immunofluorescent staining was used to assess the densities of CD8^+^, CD20^+^, FoxP3^+^ and CD68^+^ cells in the intraepithelial and intrastromal compartments as described in (8).

### Digital Pathology

Slides were scanned using an Evident VS200 whole slide image (WSI) scanner (Evident, Tokyo, Japan) and reviewed by two pathologists (KB, VHK). Image analysis was carried out using HALO digital image analysis software (Indica Labs, Corrales, NM, USA). The TMA slides were de-arrayed into individual spot images of each tissue sample and linked to clinical annotations. To localise and quantify tumour and stroma tissue, a deep neural network algorithm was trained using pathologists annotations, based on a pretrained DenseNet AI v2 network. Necrosis, erythrocyte aggregates, folds, blurry regions and glass background were excluded from further analysis. Graphical overlays were generated for each tissue class and the classification accuracy was reviewed. The total area of each tissue class was quantified in mm^2^. For nucleus segmentation, we used the HALO AI pre-trained convolutional neural network which we fine-tuned with application specific training examples. imCMS calls were assigned by digital pathology using the algorithms published in (27,28) and assigned on WSI level. imCMS was called per whole-slide image. Image analysis and marker quantification were performed blinded to clinical outcome.

The tumour stroma ratio (TSR) can easily be assessed on haematoxylin and eosin (H&E)-stained slides, and automated assessment is possible (29). The TSR is defined as the proportion of stromal area within the tumour, calculated as *stroma area/(stroma+tumour area)* (29). Previous research recommends calculating TSR in the most invasive region of the primary tumour; we analysed the TSR in both the Tumour Centre (TC) and the Invasive Margin (IM) (30).

For our analysis marker expression was defined as ANXA1 and LGR5 positive area as a percentage of total core area respectively. Dual positive area was included in ANXA1 and LGR5 area percentages. Dual positive areas made up 0.14% of classified area on average compared with 1.75% average ANXA1 positivity and 2.27% LGR5 positivity.

### Statistical analysis

Statistical analyses were conducted using R (4.5.3) within the RStudio environment (version 2026.01.1). All p-values were calculated for two-tailed tests, with statistical significance set at p < 0.05. The Kruskal–Wallis rank sum test and the Pearson chi-squared test were used to analyse the associations between the variables. Survival analyses were conducted using Kaplan–Meier (KM) estimates with the *survival* and *survminer* packages in R, and differences were assessed using log-rank tests or cox proportional hazards models. Analyses were performed separately for TC and IM samples. TC was designated as the primary sample source, as IM cores contained substantially less tumour epithelium on average.

## Results

In this study, we quantified ANXA1 and LGR5 expression using in situ hybridisation (ISH), enabling automated quantitative, spatial, compartment-driven analysis (**Figure 2**). Both CMS and the tumour-stroma ratio (TSR) were determined using a histology-based AI classifier (27,28) and correlated with histological and clinical features of disease. Our analysis was based on the evaluation of 3899 tissue samples. Of these, 2011 obtained from the tumour centre (TC) and 1888 from the invasive margin (IM).

**Figure 2.**
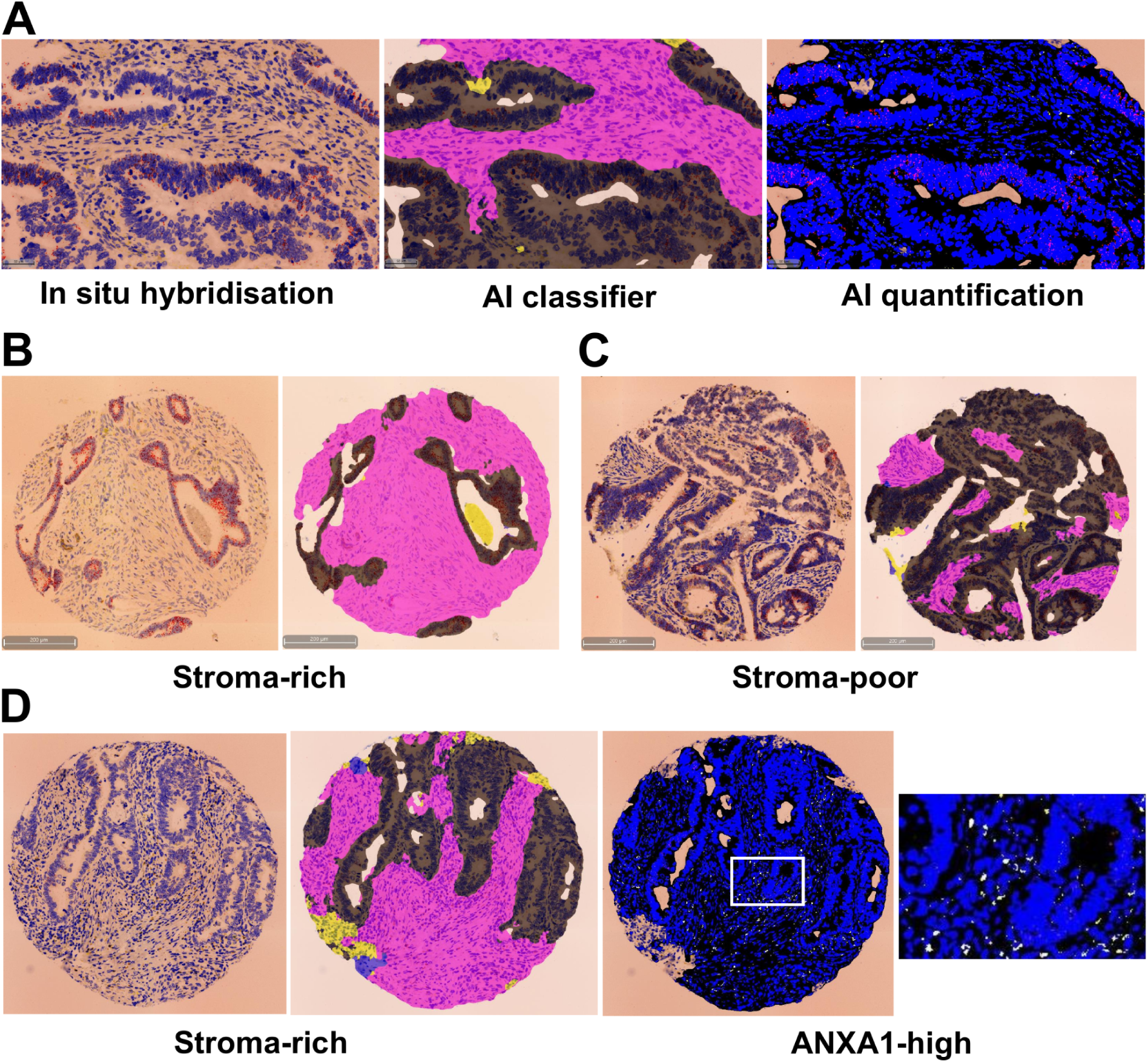
Digital quantification of stem cell markers ANXA1 and LGR5 using in-situ hybridisation. **(A)** In-situ hybridisation (ISH), Artificial Intelligence (AI)-based classification of compartments (pink: stroma; black: tumour; yellow: necrosis; blue: artifact) and compartment-based quantification of marker expression (ANXA1: yellow; LGR5: red). **(B)** Stroma-rich tissue core (left: ISH; right: compartments). **(C)** Stroma-poor tissue core. **(D)** Stroma-rich tissue core showing abundant stromal ANXA1 signal in ISH. *Scale bars* as indicated.

### Stromal content varies among Consensus Molecular Subtypes

In CRC, a high TSR is often associated with CMS4 and linked to a poor prognosis (31). Here, we assessed the correlation between the TSR and CMS subtype, with differences between groups being most pronounced in the TC samples in our dataset (**Figure 3**). As expected, we found CMS4 to have the highest median TSR (median 0.50, IQR 0.42, n = 466), followed by CMS1 (median 0.36, IQR 0.45, n = 488) and CMS3 (median 0.29, IQR 0.31, n = 272). CMS2 had the lowest median TSR (median 0.25, IQR 0.35, n = 725). Overall, we observed highly significant differences between groups (Kruskal-Wallis, p<0.001).

**Figure 3.**
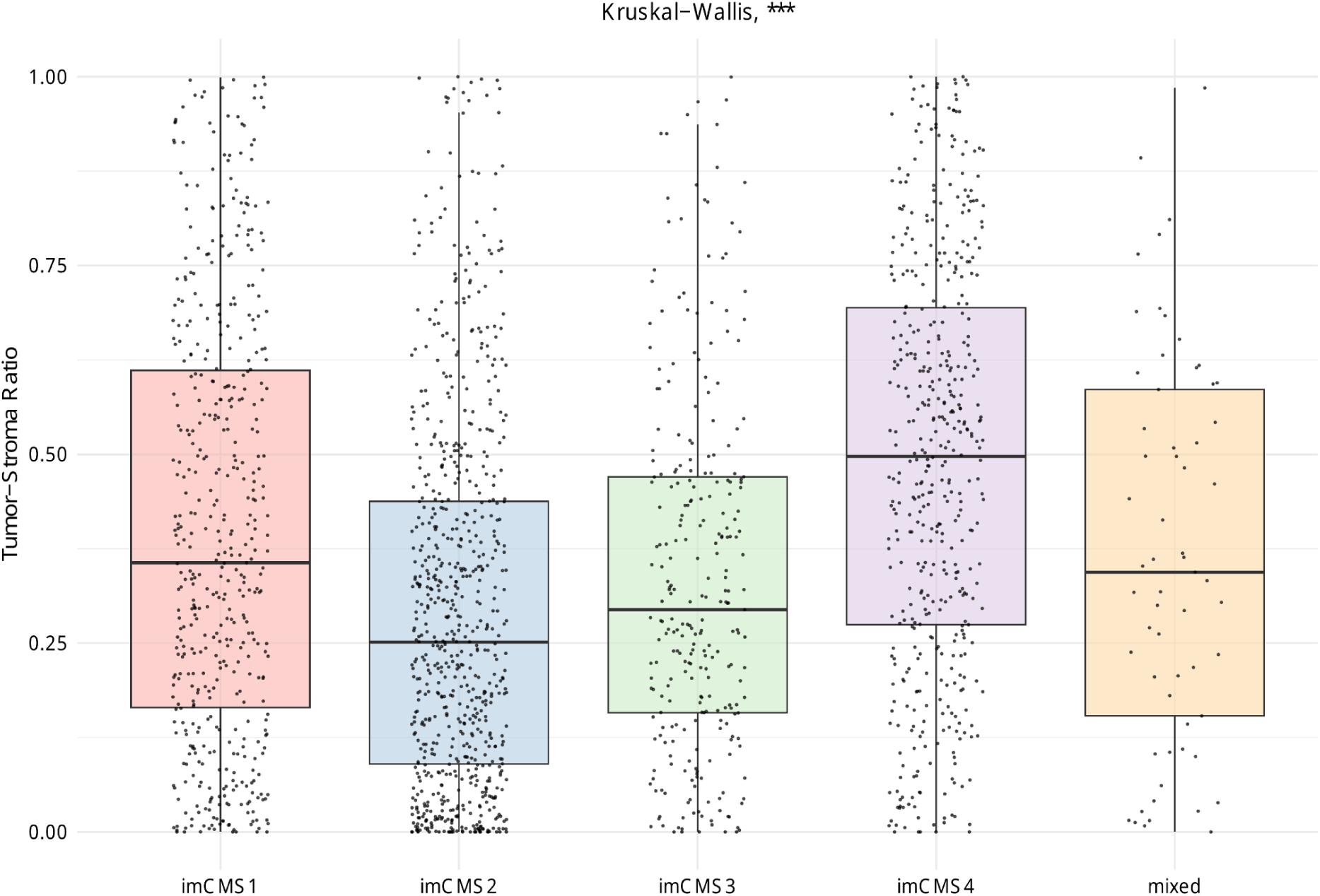
Tumour-Stroma Ratio (TSR) by image based consensus molecular subtype (imCMS). Values determined by division of stromal area by the sum of stroma and tumour epithelium areas. Data obtained from 2011 TC samples. Statistical significance was calculated using the Kruskal-Wallis test with following significance levels: ns = not significant (p > 0.05), * = p < 0.05, ** = p < 0.01, *** = p < 0.001.

### ANXA1 and LGR5 expression patterns differ among CMS

To assess the suitability of ANXA1 and LGR5 as markers for (opposing) stem cell states and to determine whether these markers are indeed associated with opposing clinicopathological characteristics, we first examined each marker individually. First, we investigated ANXA1 and LGR5 expression by CMS subgroup. For this analysis we included all samples regardless of localisation. We quantified ANXA1 and LGR5 expression over the entire tissue core (**Figure 4**), and separately within the tumour and stroma compartment.

**Figure 4.**
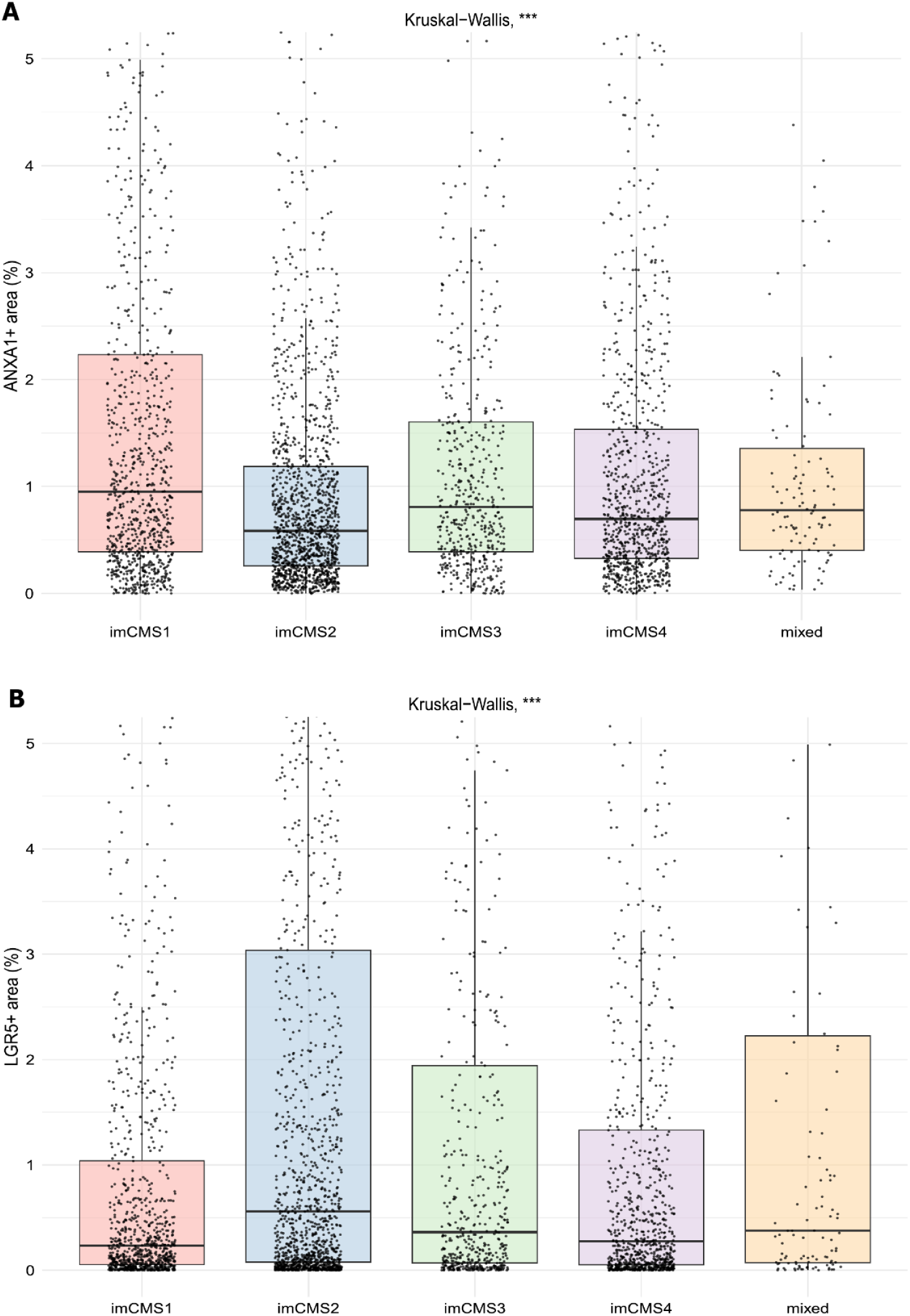
ANXA1 and LGR5 expression levels by CMS. **(A)** In-situ hybridisation (ISH) of ANXA1 on tissue samples shows marker expression patterns by image based consensus molecular subtype (CMS). Scale percentage (%) of the total core area. All available samples in our dataset were included regardless of sampling area (TC and IM). The total number of samples is 3899. **(B)** ISH of LGR5 on tissue samples shows marker expression patterns by image based consensus molecular subtype (CMS). Scale percentage (%), of the total core area. All available samples in our dataset were included regardless of sampling area (TC and IM). The total number of samples is 3899. Statistical significance was calculated using the Kruskal-Wallis test with following significance levels: ns = not significant (p > 0.05), * = p < 0.05, ** = p < 0.01, *** = p < 0.001.

Assessment of ANXA1 and LGR5 differentiated by CMS revealed opposing associations. ANXA1 expression differed significantly among CMS subgroups (Kruskal-Wallis, p<0.001). ANXA1 was most highly expressed in CMS1 cases (median 0.95, IQR 1.85, n = 953), while being comparatively suppressed in CMS2 (median 0.59, IQR 0.93, n = 1418). CMS3 (median 0.79, IQR 1.21, n = 524) and CMS4 (median 0.70, IQR 1.22, n = 892) showed intermediate expression of ANXA1. Conversely, LGR5 was most highly expressed in CMS2 cases (median 0.56, IQR 2.97, n = 1418), while being the lowest in CMS1 (median 0.24, IQR 1.00, n = 953). CMS3 (median 0.37, IQR 1.88, n = 524) and CMS4 (median 0.27, IQR 1.28, n = 892) displayed intermediate expression.

Compartment analysis (**Supplementary Tables S1A-D**) showed relative absence of LGR5 in the stromal compartment among all CMS (medians 0.08–0.09). ANXA1 was uniformly elevated (medians 0.86–0.99). In the tumour compartment, CMS1 was the only CMS subtype to have higher median ANXA1 than LGR5 expression (ANXA1: median 0.77, IQR 2.68, n = 953; LGR5: median 0.44, IQR 2.17, n = 953). The ratio of ANXA1+ to LGR5+ area in the tumour compartment was similarly significantly elevated in CMS1 in comparison to all other CMS groups (**Supplementary Figure S1**). This is consistent with RSC dominance in CMS1, in contrast to CBC dominance in the tumour compartments of CMS2, CMS3 and CMS4.

ANXA1 and LGR5, showing compartment specific behavior as described above, were correlated with stromal content. The ratio of ANXA1 to LGR5 area was correlated with tumour-stroma ratio (Pearson’s correlation, r = 0.54, p<0.001) (**Supplementary Figure S2A**). This correlation was similar across all 4 CMS subtypes showing strong dispersion, suggesting such correlation is independent from biological phenotype and microenvironment (r = 0.46, 0.61, 0.52, 0.57 for CMS1-4 respectively) (**Supplementary Figure S2B**).

### ANXA1-dominant CRC correlates with right-sidedness, MMR deficiency and a macrophage-enriched environment

Having assessed ANXA1 and LGR5 expression levels in different CMS, we investigated ANXA1 and LGR5 expression levels in relation to patient demographics, mutational status and immune microenvironment composition. ANXA1 and LGR5 expression levels were divided into quartiles, with Q4 containing the samples with the highest ANXA1/LGR5 expression. Findings for the tumour centre (TC) are presented below, respective invasive margin (IM) results can be viewed in the supplementary file.

Overall we found high ANXA1 expression levels were associated with right-sided CRC (TC: p<0.001, IM: p<0.001), lower z-scores for stromal FoxP3 (TC: p=0.004, IM: p=0.004), and higher z-scores for epithelial CD68 (TC: p<0.001, IM: p<0.001). (**Table 1, Supplementary table S2**). Additionally, patients with MMR deficient (MMRd) CRC were significantly more common in the ANXA1-high quartile (TC: p<0.001, IM: p=0.035). In accordance with our earlier findings, CMS1 CRC was overrepresented in the highest ANXA1 quartile, while CMS2 CRC displayed a tendency towards the lowest ANXA1 quartile, with overall difference between groups being highly significant (TC: p<0.001, IM: p<0.001). TSR varied significantly between quartiles (TC: p<0.001, IM: p<0.001), with Q4 revealing the largest stromal proportion. CMS4, which is defined by stromal features, showed only modest association with ANXA1 (TC: Q1=21%, Q2=24%, Q3=29%, Q4=25% of CMS4 cases) despite elevated TSR in Q4, indicating a subgroup that is stroma rich at the histological level, yet distinct from CMS4.

When regarding LGR5 expression (**Table 2, Supplementary Table S3**), we observed the opposite tendencies, with LGR5-high CRC being associated with lower stromal content, higher FoxP3 z-scores, as well as higher rates of MMR proficiency. Additionally, LGR5-high cases were less likely to be BRAF-V600E mutated.

**Table 2.** LGR5 expression in the tumour centre (TC) and clinical parameters. ANXA1, Annexin A1; CAPOX, capecitabine+oxaliplatin; CD, cluster of differentiation; FOLFOX, Folinic Acid+Fluorouracil (5-FU)+Oxaliplatin; IM, invasive margin; imCMS, image based consensus molecular subtype; LGR5, leucine-rich repeat-containing G-protein coupled receptor 5; MMR, mismatch repair; N, number (sample size); Q, quartile; RFI, recurrence-free interval; TC, tumour centre; Tumour proportion, equals 1-TSR

| Characteristic | Q1<br>N = 503<sup>a</sup> | Q2<br>N = 503<sup>a</sup> | Q3<br>N = 502<sup>a</sup> | Q4<br>N = 503<sup>a</sup> | p value<sup>b</sup> |
| --- | --- | --- | --- | --- | --- |
| Age | 65 (58, 71) | 65 (59, 71) | 65 (59, 70) | 65 (58, 70) | >0.9 |
| **Sex** |  |  |  |  | 0.7 |
| Female | 206 (25%) | 192 (24%) | 208 (26%) | 202 (25%) |  |
| Male | 297 (25%) | 311 (26%) | 294 (24%) | 301 (25%) |  |
| Performance status | 169 (28%) | 159 (27%) | 139 (23%) | 129 (22%) | 0.024 |
| **Sidedness** |  |  |  |  | 0.6 |
| Left | 291 (25%) | 287 (24%) | 294 (25%) | 305 (26%) |  |
| Right | 205 (25%) | 211 (26%) | 204 (25%) | 191 (24%) |  |
| **Trial arm** |  |  |  |  | 0.2 |
| 12 weeks | 261 (26%) | 233 (23%) | 250 (25%) | 261 (26%) |  |
| 24 weeks | 242 (24%) | 270 (27%) | 252 (25%) | 242 (24%) |  |
| **Planned treatment** |  |  |  |  | 0.062 |
| CAPOX | 364 (26%) | 363 (26%) | 364 (26%) | 332 (23%) |  |
| FOLFOX | 139 (24%) | 140 (24%) | 138 (23%) | 171 (29%) |  |
| T4 stage | 181 (27%) | 174 (26%) | 178 (26%) | 139 (21%) | 0.016 |
| **Nodal stage** |  |  |  |  | 0.8 |
| 0 | 110 (27%) | 100 (25%) | 107 (26%) | 90 (22%) |  |
| 1 | 265 (24%) | 277 (25%) | 271 (25%) | 287 (26%) |  |
| 2 | 128 (25%) | 126 (25%) | 124 (25%) | 126 (25%) |  |
| Tumour proportion | 68 (41, 88) | 61 (35, 81) | 66 (44, 84) | 72 (52, 88) | <0.001 |
| CD8, intraepithelial (z-score) | 0.14 (-0.41, 0.69) | 0.14 (-0.38, 0.67) | 0.12 (-0.50, 0.55) | 0.20 (-0.36, 0.70) | 0.2 |
| CD8, intrastromal (z-score) | 0.12 (-0.57, 0.66) | 0.15 (-0.43, 0.67) | 0.06 (-0.55, 0.62) | 0.13 (-0.50, 0.61) | 0.4 |
| FOXP3, intraepithelial (z-score) | 0.20 (-0.28, 0.62) | 0.18 (-0.29, 0.56) | 0.13 (-0.40, 0.56) | 0.27 (-0.16, 0.58) | 0.013 |
| FOXP3, intrastromal (z-score) | 0.07 (-0.70, 0.62) | 0.05 (-0.60, 0.59) | 0.19 (-0.52, 0.70) | 0.38 (-0.30, 0.84) | <0.001 |
| CD20, intraepithelial (z-score) | 0.20 (-0.52, 0.69) | 0.20 (-0.62, 0.73) | 0.20 (-0.54, 0.67) | 0.22 (-0.42, 0.72) | 0.7 |
| CD20, intrastromal (z-score) | 0.10 (-0.47, 0.58) | 0.23 (-0.38, 0.69) | 0.07 (-0.49, 0.61) | 0.11 (-0.45, 0.60) | 0.10 |
| CD68, intraepithelial (z-score) | -0.01 (-0.62, 0.66) | 0.06 (-0.51, 0.65) | 0.11 (-0.49, 0.77) | -0.01 (-0.48, 0.68) | 0.2 |
| CD68, intrastromal (z-score) | 0.06 (-0.45, 0.51) | 0.09 (-0.34, 0.50) | 0.08 (-0.32, 0.50) | 0.15 (-0.37, 0.53) | 0.6 |
| **FOXP3 (intrastromal) / CD8 (intraepithelial)** |  |  |  |  | 0.006 |
| high/high | 143 (22%) | 144 (23%) | 165 (26%) | 186 (29%) |  |
| high/low | 195 (26%) | 187 (25%) | 168 (23%) | 188 (25%) |  |
| low/low | 164 (26%) | 170 (27%) | 169 (27%) | 127 (20%) |  |
| **imCMS** |  |  |  |  | <0.001 |
| imCMS1 | 150 (31%) | 148 (30%) | 112 (23%) | 78 (16%) |  |
| imCMS2 | 157 (22%) | 150 (21%) | 186 (26%) | 232 (32%) |  |
| imCMS3 | 73 (27%) | 64 (24%) | 62 (23%) | 73 (27%) |  |
| imCMS4 | 110 (24%) | 128 (27%) | 129 (28%) | 99 (21%) |  |
| Mixed | 13 (22%) | 13 (22%) | 13 (22%) | 21 (35%) |  |
| **MMR status** |  |  |  |  | <0.001 |
| Deficient | 82 (39%) | 64 (31%) | 43 (21%) | 19 (9.1%) |  |
| Proficient | 342 (22%) | 358 (23%) | 404 (26%) | 434 (28%) |  |
| **BRAF V600E** |  |  |  |  | 0.034 |
| Mutant | 15 (30%) | 16 (32%) | 13 (26%) | 6 (12%) |  |
| Wild-type | 87 (24%) | 88 (24%) | 75 (20%) | 119 (32%) |  |
| **KRAS** |  |  |  |  | 0.5 |
| Mutant | 36 (26%) | 28 (20%) | 30 (22%) | 45 (32%) |  |
| Wild-type | 66 (24%) | 76 (27%) | 58 (21%) | 80 (29%) |  |

<sup>a</sup>Median (Q1, Q3); n (%).
<sup>b</sup>Kruskal–Wallis rank sum test; Pearson’s chi-squared test.

### ANXA1 and LGR5 are associated with opposing clinicopathological features and represent a phenotypic spectrum

As we observed that ANXA1 and LGR5 behaved inversely and were associated with opposing CMS, we hypothesised that the relationship between ANXA1 and LGR5 would be more consequential for clinicopathological features than the individual markers taken by themselves. Therefore we decided to combine ANXA1 and LGR5 into a single measure. We implemented the ANXA1-LGR5 measure by subtracting the LGR5 area from the ANXA1 area and dividing the values into four quartiles.

We found the middle two quartiles to contain relatively equal levels of ANXA1 and LGR5 expression, whereas Q4 and Q1 showed a clear dominance of either ANXA1 or LGR5, and which correspond to RSC and CBC dominant cancer in our framework **(Supplementary Figures S3A, S3B)**.

We repeated the analysis with our combined ANXA1-LGR5 measure (**Table 3, Supplementary Table S4**). We see the same directional trends for sidedness, TSR, stromal FoxP3, epithelial CD68, CMS and MMR loss. The combined ANXA1-LGR5 measure additionally showed more uniform gradients for MMR-deficiency and CMS1. The association with BRAF-V600E was also stronger than either marker alone (TC: p=0.001), although this rested on only 50 mutated cases in our dataset and was not reproduced in the invasive margin (IM: p=0.3). Conversely, associations with sidedness and epithelial CD68 were weaker in the combined measure. Notably, the gradient in tumour proportion was shallower for the combined measure than for ANXA1 alone (medians 74%, 69%, 68%, 50% vs 84%, 69%, 58%, 51%), suggesting that subtracting LGR5 partly offsets the dependency of ANXA1 on stromal content. Metastatic sites and recurrence were assessed only for the combined measure, and showed that peritoneal metastasis was more frequent in the ANXA1-dominant quartile (TC: p=0.019), with 43% of affected patients in Q4.

**Table 3.**
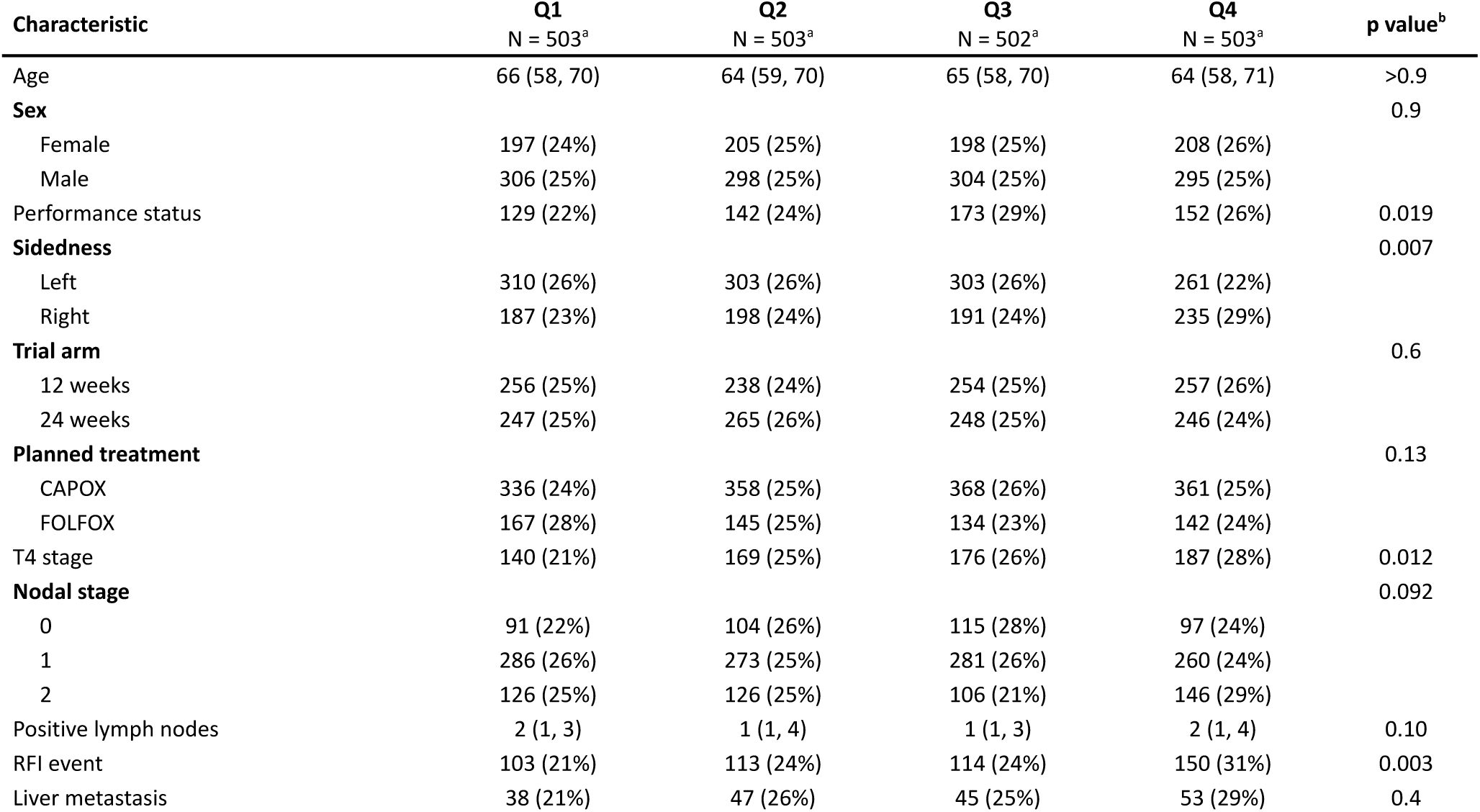
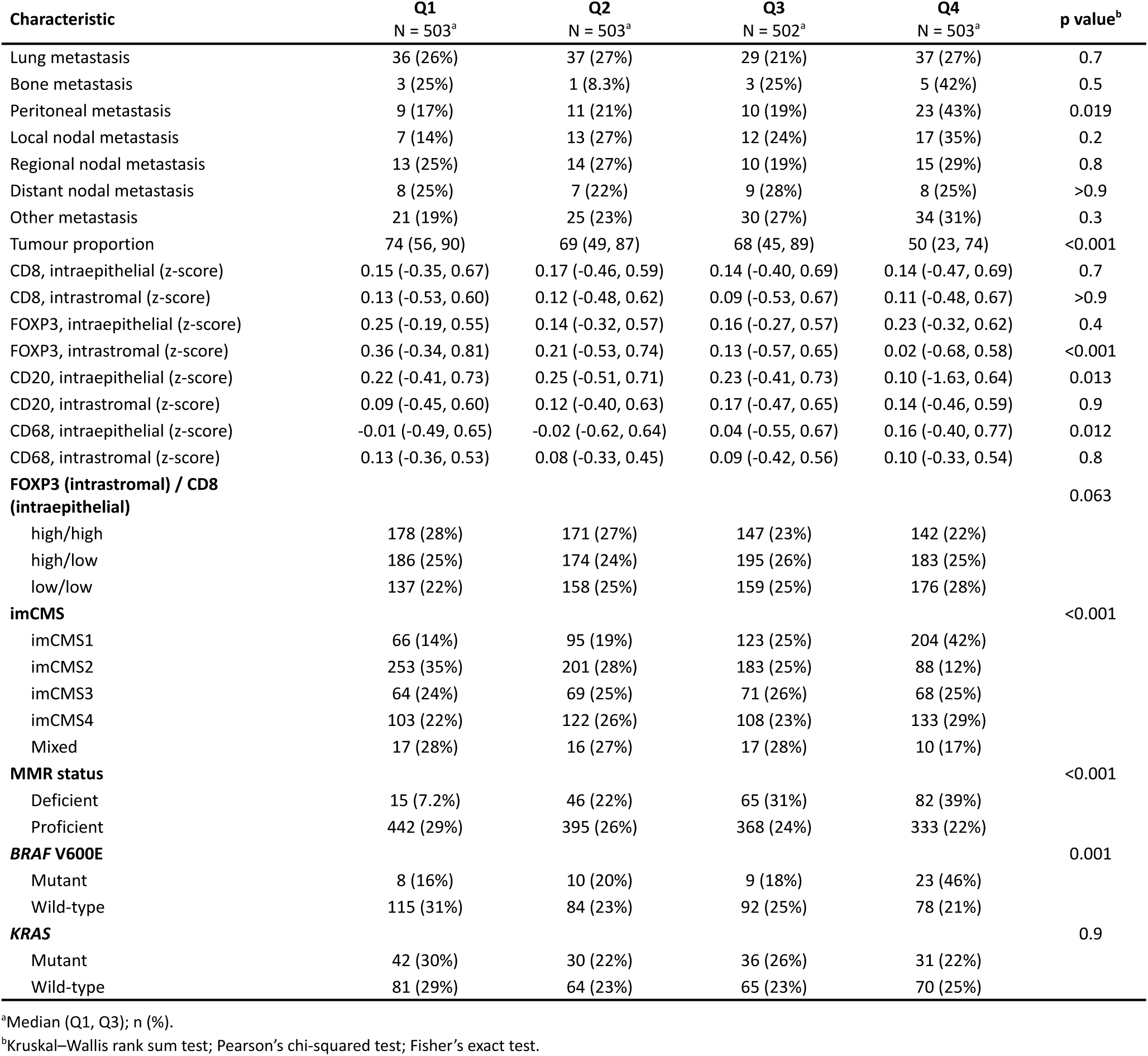
ANXA1-LGR5 quartiles in the tumour centre (TC) and clinical data. ANXA1, Annexin A1; CAPOX, capecitabine+oxaliplatin; CD, cluster of differentiation; FOLFOX, Folinic Acid+Fluorouracil (5-FU)+Oxaliplatin; IM, invasive margin; imCMS, image based consensus molecular subtype; LGR5, leucine-rich repeat-containing G-protein coupled receptor 5; MMR, mismatch repair; N, number (sample size); Q, quartile; RFI, recurrence-free interval; TC, tumour centre; Tumour proportion, equals 1-TSR

### Worse outcome in ANXA1-dominant CRC

In the TC, combining ANXA1 and LGR5 into a single measure showed better prognostic performance than both individual ANXA1 and LGR5 levels, despite LGR5 not being an independent prognostic marker on its own **(Figure 5A)**. Individual ANXA1 expression in TC samples significantly affected overall survival (OS) (p=0.043), cancer specific survival (CSS) (p=0.02) and recurrence free interval (RFI) (p=0.017), with high ANXA1 being associated with worse survival measures (**Figure 5B**). LGR5 expression on the other hand was not significantly associated with any of the outcome measures (**Figure 5C**). Integrating ANXA1 and LGR5 substantially improved prognostic stratification between groups for TC samples, with the ANXA1 dominant (RSC) quartile demonstrated markedly worse CSS and RFI (CSS: p<0.001, RFI: p=0.004), while OS stratification remained similar (p=0.044). In Invasive Margin samples, we generally see reduced prognostic stratification by using IM samples when compared to TC samples (**Supplementary Figures S4A, S4B, S4C**). Restricting analysis to CMS1 TC cases showed no significant differences between quartiles, though small subgroup size limited power to detect an effect (**Supplementary Figure S5A**). Restricting analysis to CMS4 cases, showed similar directional trends to overall analysis, while significantly worse outcome was only observed for cancer specific survival (**Supplementary Figure S5B**)

**Figure 5.**
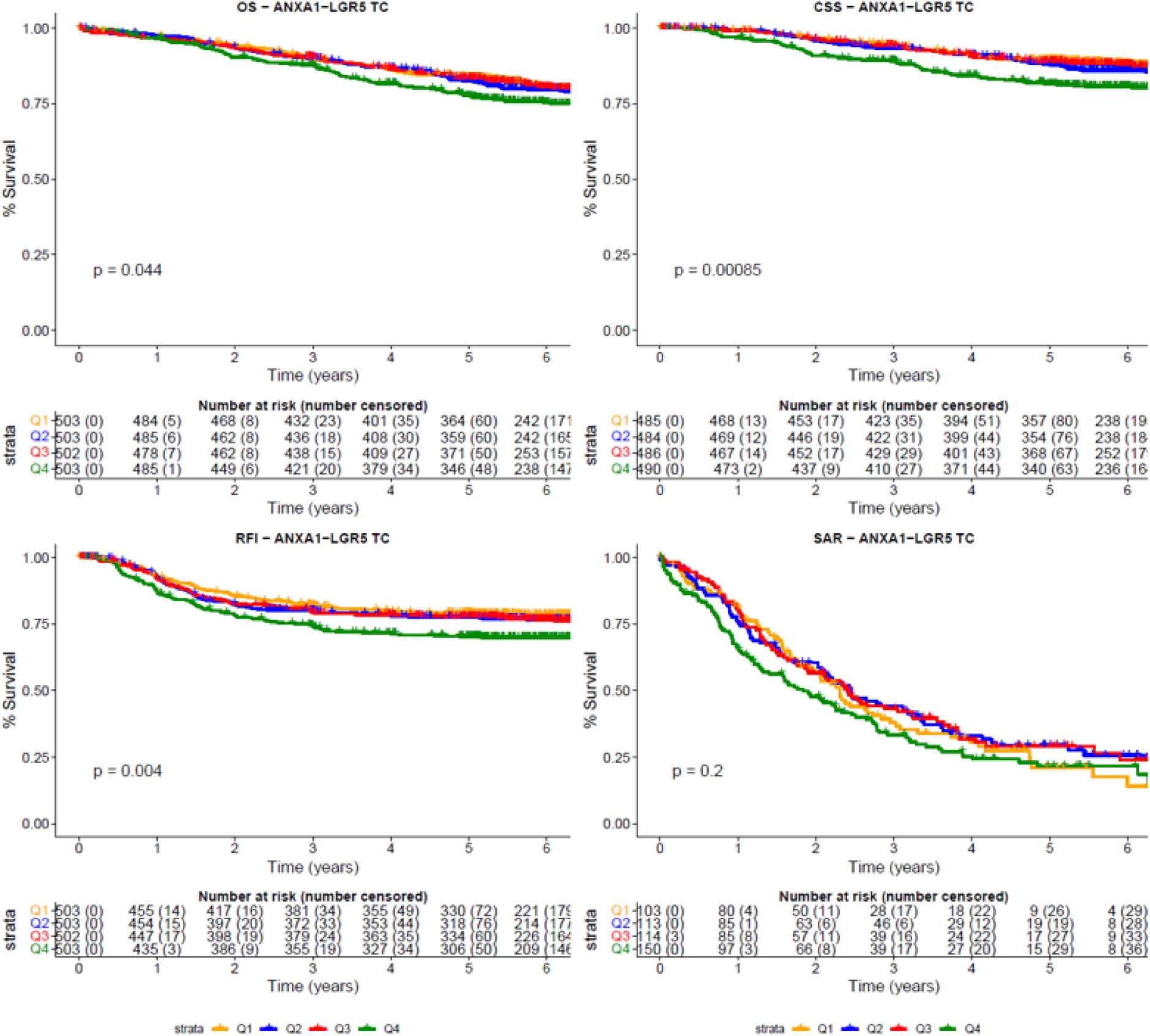
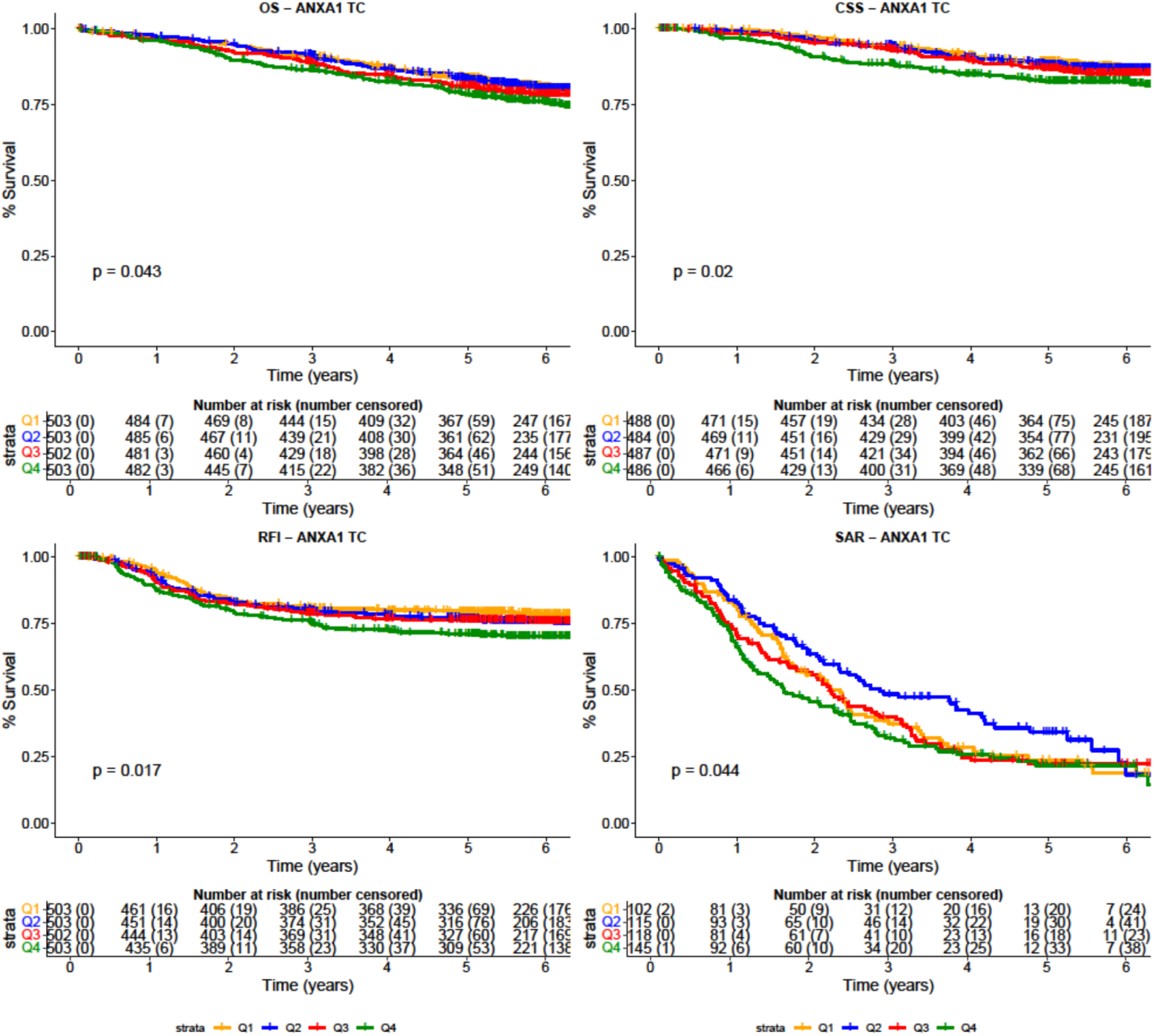
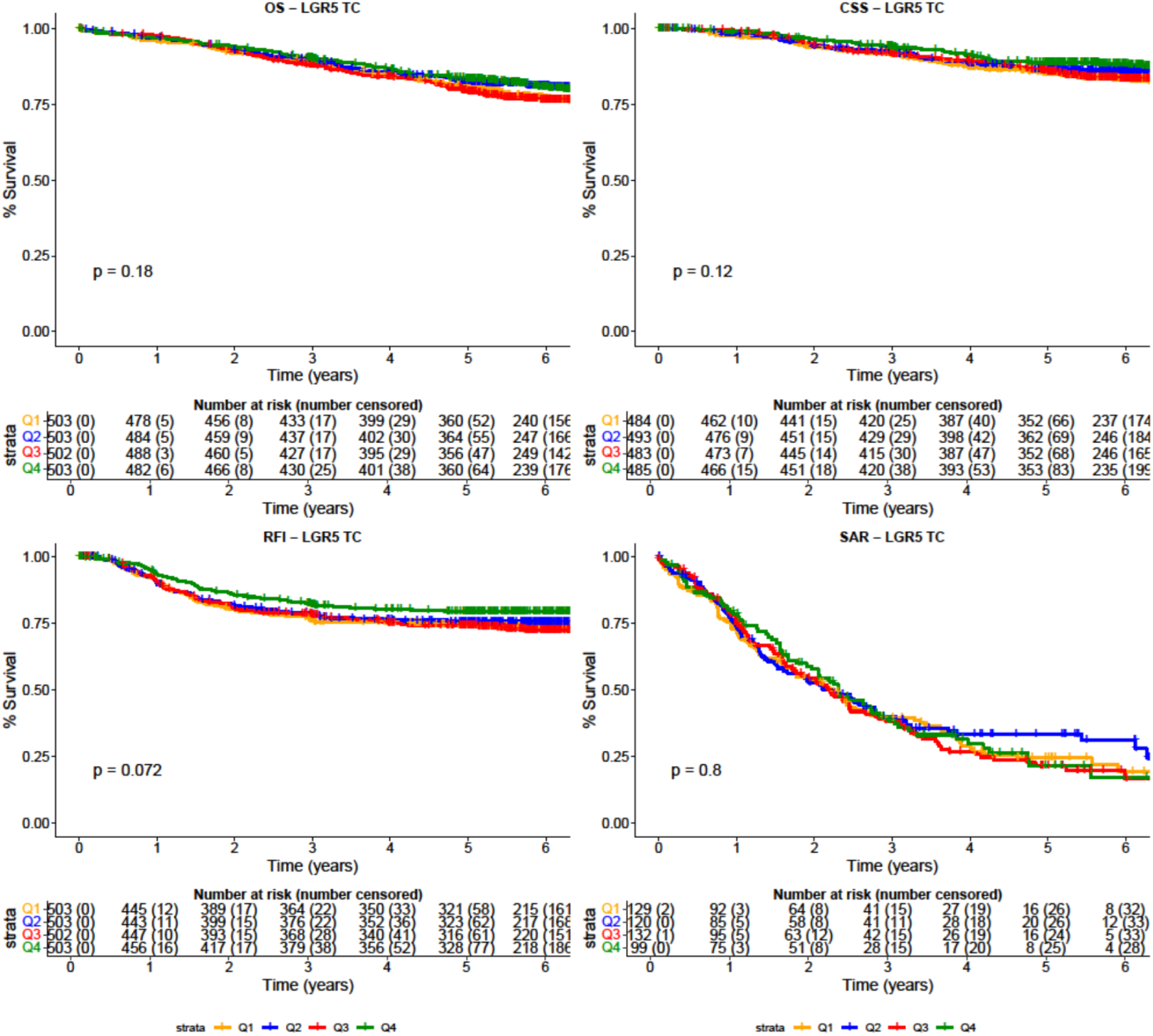
High ANXA1 signal correlates with worse outcomes. (A) Survival analysis by ANXA1-LGR5 expression quartiles in tumour centre (TC) samples. Samples have been ranked by subtracting the LGR5 area from the ANXA1 area (%). Q4 contains cases with highest relative ANXA1 expression, Q1 contains cases with highest relative LGR5 expression. P-values determined by log-rank test. *CSS, cancer specific survival; OS, overall survival; RFI, recurrence free interval; SAR, survival after relapse* (B) Survival analysis by ANXA1 expression quartiles in TC samples. Samples have been ranked by ANXA1 positive area (%). Q4 contains cases with highest ANXA1 expression, Q1 contains cases with lowest ANXA1 expression. P-values determined by log-rank test. *CSS, cancer specific survival; OS, overall survival; RFI, recurrence free interval; SAR, survival after relapse* (C) Survival analysis by LGR5 expression quartiles in TC samples. Samples have been ranked by LGR5 positive area (%). Q4 contains cases with highest LGR5 expression, Q1 contains cases with lowest LGR5 expression. P-values determined by log-rank test. *CSS, cancer specific survival; OS, overall survival; RFI, recurrence free interval; SAR, survival after relapse*.

Given the poor prognosis of ANXA1 dominant CRC, we investigated whether this subgroup derived additional benefit from longer treatment. Within our ANXA1-dominant TC samples, patients randomised to 3 months vs. 6 months showed no difference in outcome (**Figure 6**). This indicates no evidence of differential benefit for ANXA1 dominance and is in line with the overall noninferiority of 3 months of chemotherapy in the SCOT Trial.

**Figure 6.**
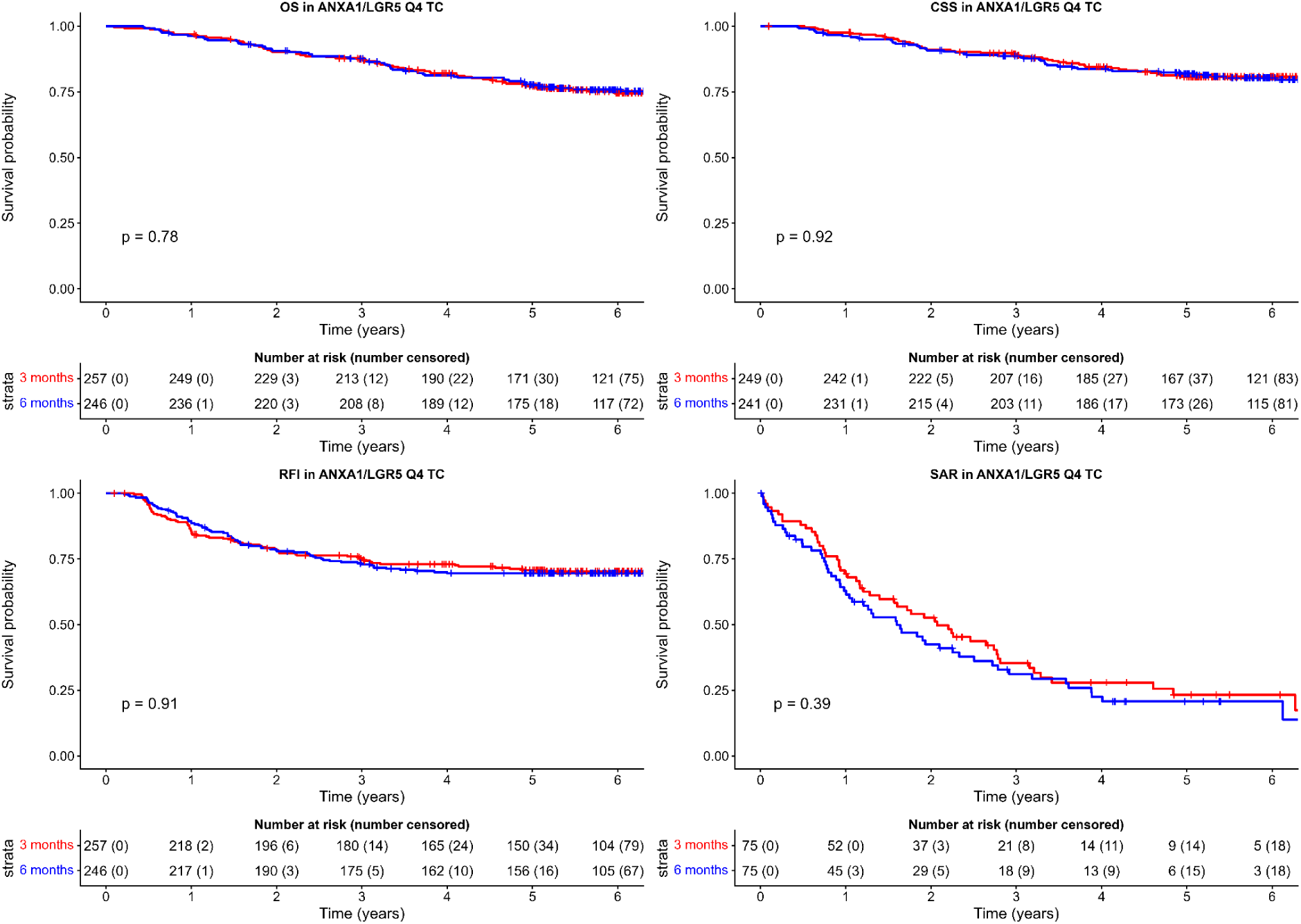
ANXA1 dominant cases show no benefit from prolonged chemotherapy. Kaplan–Meier survival analysis by treatment arm in ANXA1-dominant colorectal cancer cases. Cases were ranked by subtracting the LGR5 area from the ANXA1 area (%) in the tumour centre and divided into quartiles. Only Q4, containing cases with the highest relative ANXA1 expression, is shown. Numbers at risk (censored) are shown beneath each panel; vertical ticks denote censored observations. P-values determined by log-rank test. *CSS, cancer-specific survival; OS, overall survival; RFI, recurrence-free interval; SAR, survival after relapse; TC, tumour centre*.

In order to assess independence of our ANXA1-LGR5 measure from additional prognostic factors, we performed a multivariate cox regression, adjusting for TSR, T-stage, N-stage, sidedness, treatment arm and type, sex, age and MMR status (**Table 4, Supplementary Table S5**). In the multivariate analysis of TC samples we observed a collapse of OS signal (Q4 vs Q1: HR 1.09, 95% CI 0.79-1.49, p=0.599), with stage, sidedness and age being the dominant prognostic factors. Considering CSS, the effect magnitude remained relevant, but not statistically significant (Q4 vs Q1: HR 1.46, 95% CI 0.98-2.16, p=0.061). For RFI however, we observed a statistically significant signal between Q1 and Q4 despite adjustment for confounding factors, with considerable estimated effect size (Q4 vs Q1: HR 1.43, 95% CI 1.08-1.90, p=0.014). Critically, this effect was independent of MMR status which remained strongly protective (RFI: HR 0.50, 95% CI 0.34-0.73, p<0.001). This indicates ANXA1-dominance in the tumour centre as an independent recurrence-risk factor.

**Table 4:** Multivariable analysis for tumour centre (TC) samples. *FOLFOX*: Folinic Acid+Fluorouracil (5-FU)+Oxaliplatin; *M*: Male; *MMR*: mismatch repair; *N*: nodal; *Q*: quartile. *Tumour proportion as a continuous variable

| Variable | OS (N=1727) |  | CSS (N=1666) |  | RFI (N=1727) |  | SAR (N=416) |  |
| --- | --- | --- | --- | --- | --- | --- | --- | --- |
|  | HR (CIs) | p | HR (CIs) | p | HR (CIs) | p | HR (CIs) | p |
| ANXA1-LGR5 Q2 vs Q1 (ref) | 0.93 (0.68-1.26) | 0.619 | 1.09 (0.73-1.62) | 0.679 | 1.01 (0.76-1.35) | 0.928 | 0.82 (0.58-1.16) | 0.266 |
| ANXA1-LGR5 Q3 vs Q1 (ref) | 0.89 (0.65-1.22) | 0.452 | 0.92 (0.60-1.40) | 0.686 | 1.09 (0.81-1.45) | 0.571 | 0.74 (0.52-1.06) | 0.105 |
| ANXA1-LGR5 Q4 vs Q1 (ref) | 1.09 (0.79-1.49) | 0.599 | 1.46 (0.98-2.16) | 0.061 | 1.43 (1.08-1.90) | 0.014 | 0.83 (0.59-1.16) | 0.270 |
| Tumour proportion* | 1.00 (0.99-1.00) | 0.0397 | 1.00 (0.99-1.00) | 0.085 | 1.00 (0.99-1.00) | 0.033 | 1.00 (0.99-1.00) | 0.105 |
| T4 stage | 2.00 (1.61-2.49) | <0.001 | 2.45 (1.86-3.22) | <0.001 | 2.05 (1.68-2.50) | <0.001 | 1.33 (1.05-1.70) | 0.021 |
| N stage | 1.67 (1.42-1.96) | <0.001 | 2.05 (1.66-2.51) | <0.001 | 1.72 (1.49-2.00) | <0.001 | 1.32 (1.09-1.59) | 0.005 |
| Sidedness Right | 1.55 (1.23-1.94) | <0.001 | 1.94 (1.46-2.57) | <0.001 | 1.37 (1.12-1.69) | 0.003 | 1.80 (1.40-2.33) | <0.001 |
| Arm 24 weeks | 0.92 (0.74-1.13) | 0.423 | 0.80 (0.61-1.05) | 0.103 | 0.89 (0.73-1.08) | 0.229 | 1.15 (0.90-1.46) | 0.260 |
| Planned treatment Folfox | 0.95 (0.76-1.20) | 0.693 | 0.91 (0.68-1.22) | 0.523 | 1.05 (0.85-1.29) | 0.661 | 0.87 (0.67-1.13) | 0.297 |
| Sex M | 0.97 (0.78-1.21) | 0.788 | 0.74 (0.57-0.97) | 0.031 | 0.91 (0.75-1.11) | 0.348 | 0.79 (0.62-1.02) | 0.068 |
| Age at registration | 1.02 (1.01-1.03) | 0.006 | 1.01 (0.99-1.02) | 0.299 | 0.99 (0.98-1.00) | 0.235 | 1.03 (1.01-1.04) | <0.001 |
| MMR IHC loss | 0.71 (0.49-1.03) | 0.070 | 0.58 (0.36-0.92) | 0.022 | 0.50 (0.34-0.73) | <0.001 | 1.34 (0.86-2.09) | 0.199 |

## Discussion

Our spatially resolved analysis of ANXA1 and LGR5 in TMAs of the SCOT trial revealed differences in the expression of stem cell markers across spatial compartments and molecular subtypes. As expected, our AI classifier detected the highest stromal proportion in CMS4. However, CMS4 did not show a strong tendency towards either ANXA1 or LGR5 expression, which is notable given the strong association between ANXA1 and TSR. Instead, ANXA1 was found to be primarily associated with CMS1, while LGR5 was found to be highly associated with CMS2. The ambivalent stem cell marker expression of CMS4 corresponds to the refined CMS classification (5), which states that CMS4 is determined by stromal features, these however being independent of the epithelial programme. Indeed, in CMS4 we find compartment specific ANXA1/LGR5 expression signatures. This stands in contrast to CMS1 where ANXA1 outweighs LGR5 expression in both the tumour and stroma compartments. The expression of ANXA1 and other RSC markers by stromal and immune cells could explain why bulk transcriptomic data found CMS4 to appear to lean towards the RSC phenotype. Therefore, our spatially resolved analysis could help reconcile the conflicting findings of earlier research (15). Similarly, in CMS1 the observed ANXA1 signal in the tumour compartment could stem from the CD68^+^ macrophage infiltration evident in our data in addition to being expressed by the tumour cells themselves. ANXA1 is considered an important immunoregulatory protein, with promotion of M2-like macrophage polarisation (23). Therefore high levels of ANXA1 could contribute to immune suppression and evasion in normally immunogenic cancer, explaining the especially strong associations found in the CMS1 group.

We observe ANXA1-dominant CRC to be associated with right-sidedness, MMRd, and increased occurrence of BRAF mutations, features which are characteristic of CMS1 (4). MMR deficiency is associated with lower stromal content overall (32). Notably, we find our ANXA1 dominant group to be simultaneously high in stromal content and with high rates of MMR-deficiency. Other research has also independently identified an MMR-deficient group of CRC with high stromal content which is associated with treatment resistance and worse outcome (33), which could correspond to our ANXA1-dominant group. Our research suggests ANXA1-dominance to be associated with more advanced stages of CRC, with higher rates of T4-stage and peritoneal metastasis. Despite the association with progressed disease, we nonetheless found ANXA1-dominant CRC in TC samples to be a negative recurrence risk predictor independent of T4-stage status, nodal status, age, TSR, and MMR status.

ANXA1 was first described as an effector of glucocorticoid signalling and as a mediator of inflammation suppression (22). We find ANXA1 to be associated with distinct TIME changes, most importantly reduced stromal FoxP3 and increased epithelial CD68 densities. Low FoxP3 levels are associated with worse outcomes in CRC in contrast to other forms of cancer. While we did not measure macrophage polarisation, studies have indicated that ANXA1/FPR2 signaling promotes macrophage polarization towards an M2 and inflammation resolving phenotype (23,34). Therefore ANXA1 upregulation could be an important mechanism for immune-evasion in this highly MMR-deficient group.

Stem cell plasticity in CRC has become a focus of research. Whereas physiological intestinal homeostasis is maintained by LGR5^+^ stem cells within the crypt base, cancer stem cells (CSCs) have been shown to dynamically adapt to environmental cues and cellular stress (15). Notably, experimental models suggest that metastatic seeding can be mediated by functionally plastic LGR5^−^ populations (35). This CSC plasticity is thought to contribute to therapy resistance, and activation of regenerative or stress-associated programs, including ANXA1-linked signalling, has been associated with reduced chemotherapy response (36). While we find that ANXA1 dominance is associated with poor survival, we see no protective effect of longer chemotherapy. Also, previous research has demonstrated early liver metastasis to be transcriptionally homogeneous with high MAPK signaling and high RSC marker expression (37). We find ANXA1-dominant CRC to be associated with advanced disease, with higher rates of T4 and peritoneal metastasis. The specific TIME changes observed in our study, as well as higher TSR could help explain the metastatic potential of the MMR-deficient ANXA1-dominant group. Specific targeting of CSC populations is being investigated as a therapeutic measure and targeted MAPK inhibition, as well as preventing LGR5^+^ CSC reestablishment have already been proposed (38,39).

Increasing interest has focused on the role of the microbiome in the development of (early-onset) CRC (40,41). F. nucleatum, a Gram-negative oral commensal, was demonstrated to accelerate CRC by inducing ANXA1 expression (42,43). Interestingly, we found ANXA1 dominant CRC to display similar associations as those found for F. nucleatum-enriched CRC, namely CMS1, right-sidedness and MMR deficiency (44–46). Given that ANXA1 has already been proposed as an effector of F. nucleatum driven CRC and that F. nucleatum has furthermore been associated with Ly6a mediated RSC activation (47), the interactions between F. nucleatum, ANXA1 and CMS1 cancer warrant further investigation.

The limitations of this work are the restricted multiplex setup with two markers. Second, we used ISH as a single modality, which is not a routine technique in most pathology departments and is difficult to quantify visually and not easily translatable. Third, ISH might not fully capture the dynamic phenotypic spectrum of stem cell phenotypes in CRC. Furthermore, the SCOT Trial included only stage II high-risk as well as stage III cases, with all patients receiving the adjuvant oxaliplatin doublet. Therefore these results are not generalisable to other stages or untreated patients. On the other hand, the strengths of this paper include the large patient cohort, the large amount of parameters evaluated and the quality of the underlying SCOT trial. Furthermore, we have access to a large amount of data points, combining immune, genetic, prognostic and survival statistics in a single study.

In conclusion, we found that relative ANXA1 dominance over LGR5 in CRC was an independent risk factor for recurrence, and was associated with worse prognosis. ANXA1 was also associated with a high TSR, a higher incidence of T4 stage, peritoneal metastasis, right-sided cancer, MMRd, BRAF-V600E mutations, and a specific macrophage-rich immune microenvironment. Investigations of the role of ANXA1 in driving TIME changes as well as its role as an effector of F. nucleatum driven CRC would be exciting avenues for further investigation.

## Supporting information

REMARK checklist

## ADDITIONAL INFORMATION

### TransSCOT Consortium

The TransSCOT Trial Management Group includes (alphabetical order): David Church (Cancer Genomics and Immunology Group, The Centre for Human Genetics, University of Oxford, United Kingdom), Enric Domingo (Department of Oncology, University of Oxford, United Kingdom), Joanne Edwards (School of Cancer Sciences, University of Glasgow, Glasgow, United Kingdom), Bengt Glimelius (Uppsala University, Uppsala, Sweden), Ismail Gogenur (Centre for Surgical Science, Zealand University Hospital, Denmark), Andrea Harkin (CRUK Glasgow Clinical Trials Unit, University of Glasgow, Glasgow, United Kingdom), Jen Hay (Glasgow Tissue Research Facility, University of Glasgow, Queen Elizabeth University Hospital, Glasgow, United Kingdom), Timothy Iveson (University of Southampton, Southampton, United Kingdom), Emma Jaeger (Department of Oncology, University of Oxford, United Kingdom), Caroline Kelly (CRUK Glasgow Clinical Trials Unit, University of Glasgow, Glasgow, United Kingdom), Rachel Kerr (Department of Oncology, University of Oxford, United Kingdom), Noori Maka (Glasgow Tissue Research Facility, University of Glasgow, Queen Elizabeth University Hospital, Glasgow, United Kingdom), Hannah Morgan (Glasgow Tissue Research Facility, University of Glasgow, Queen Elizabeth University Hospital, Glasgow, United Kingdom), Karin Oien (Glasgow Tissue Research Facility, University of Glasgow, Queen Elizabeth University Hospital, Glasgow, United Kingdom), Clare Orange (NHS Greater Glasgow and Clyde Biorepository, Glasgow, United Kingdom), Claire Palles (University of Birmingham, Birmingham, United Kingdom), Campbell Roxburgh (School of Cancer Sciences, University of Glasgow, Glasgow, United Kingdom), Owen Sansom (CRUK Beatson Institute of Cancer Research, Garscube Estate, Glasgow, United Kingdom), Mark Saunders (The Christie NHS Foundation Trust, Manchester, United Kingdom), Ian Tomlinson (Department of Oncology, University of Oxford, United Kingdom).

### imCMS Consortium

The imCMS consortium includes (alphabetical order): Enric Domingo (Department of Oncology, University of Oxford, Oxford, United Kingdom), Viktor H. Koelzer (Institute of Medical Genetics and Pathology, University Hospital Basel, Basel, Switzerland; Department of Biomedical Engineering, Faculty of Medicine, University of Basel, Allschwil, Switzerland; Department of Pathology and Molecular Pathology, University Hospital Zurich, University of Zurich, Zurich, Switzerland; Nuffield Department of Medicine, University of Oxford, Roosevelt Drive, Oxford, United Kingdom), Maxime Lafarge (Institute of Medical Genetics and Pathology, University Hospital Basel, Basel, Switzerland; Department of Biomedical Engineering, Faculty of Medicine, University of Basel, Allschwil, Switzerland), Tim Maughan (Nuffield Department of Medicine, University of Oxford, Roosevelt Drive, Oxford, United Kingdom), Jens Rittscher (Big Data Institute/Li Ka Shing Center for Health Information and Discovery, University of Oxford, Oxford, United Kingdom; Ground Truth Labs Ltd, Oxford, United Kingdom; Oxford NIHR Biomedical Research Center, Oxford University Hospitals NHS Foundation Trust, Oxford, United Kingdom; Nuffield Department of Medicine, Ludwig Institute for Cancer Research, University of Oxford, Oxford, United Kingdom), Korsuk Sirinukunwattana (Big Data Institute/Li Ka Shing Center for Health Information and Discovery, University of Oxford, Oxford, United Kingdom, Ground Truth Labs Ltd, Oxford, United Kingdom, Nuffield Department of Medicine, Ludwig Institute for Cancer Research, University of Oxford, Oxford, United Kingdom, Institute of Biomedical Engineering, Department of Engineering Science, University of Oxford, Oxford, United Kingdom).

Representative imCMS Consortium: Viktor H. Koelzer.

## Ethics statement

The SCOT trial (ISRCTN59757862) was conducted in accordance with the Declaration of Helsinki and Good Clinical Practice guidelines. The trial protocol received ethical approval by the West Glasgow Research Ethics Committee (REC reference 07/S0703/136; protocol version 1.1, January 21, 2008), with all subsequent amendments approved as required. All patients provided written informed consent before enrolment, including consent for the collection of tumour tissue and its use in translational research within the TransSCOT program. The present analysis was performed on tissue collected under this consent and falls within the original ethics approval.

## Data availability

The datasets pertaining to the SCOT trial used during the current study are available from the TransSCOT collaboration on reasonable request in the absence of scientific conflict of interest. Applications for analysis of TransSCOT samples are welcome and should be addressed to JH.

## Code availability

Analyses were performed in R using publicly available packages (survival, survminer). Custom analysis scripts are available from the corresponding author upon reasonable request.

## Funding

The SCOT trial was funded by the Medical Research Council (transferred to NETSCC–Efficacy and Mechanism Evaluation; grant number G0601705), the National Institute for Health and Care Research (NIHR) Health Technology Assessment programme (14/140/84), Cancer Research UK Core Clinical Trials Unit Glasgow funding (C6716/A9894), and the Swedish Cancer Society. The TransSCOT sample collection was funded by a Cancer Research UK Clinical Trials Awards and Advisory Committee—Sample Collection award (C6716/A13941). Viktor H Koelzer acknowledges funding from the Swiss National Science Foundation (grant nos. P2SKP3_168322/1 and P2SKP3_168322/2), the ISREC Foundation and the Promedica Foundation (F-87701-41-01). The views expressed are those of the authors and not necessarily those of the National Health Service, the NIHR, or the Department of Health and Social Care. The funders had no role in the design of the study; the collection, analysis, and interpretation of the data; the writing of the manuscript; or the decision to submit the manuscript for publication.

## Competing interests

I. = immediate family member; (Inst) = payment to the author’s institution.

**Mark P Saunders**

Honoraria: Servier, Takeda, GlaxoSmithKline, Incyte, Nordic Bioscience, Bayer, MSD Travel, accommodations, expenses: Takeda, Bayer

**Joanne Edwards**

Research funding: Varian Medical Systems

**Owen J Sansom**

Research funding: Novartis, AstraZeneca, Boehringer Ingelheim, NeoPhore

**Ian Tomlinson**

Patents, royalties, other intellectual property: molecular testing panel for 5-fluorouracil toxicity

**David N Church**

Employment: Amgen (I)

Stock and other ownership interests: Amgen (I) Honoraria: GlaxoSmithKline

Research funding: HaloDx (Inst), Moderna Therapeutics (Inst), GlaxoSmithKline (Inst) Travel, accommodations, expenses: Takeda

**Viktor H Koelzer**

All of the following are unrelated to the present study. Employment: University Hospital Basel

Consulting or advisory role: advisory board, Takeda; member of the European Key Opinion Leader Network in Digital Pathology, supported by Roche

Speakers’ bureau: invited speaker for Sharing Progress in Cancer Care (a non-profit educational organisation) and Indica Labs

Research funding: sponsored research agreements with Roche and IAG

Patents, royalties, other intellectual property: participant in a patent application co-owned by the Netherlands Cancer Institute (NKI-AVL) and the University of Basel on methods for predicting treatment outcome and/or selecting a subject suitable for immune checkpoint therapy (Filing No. NL73455NL); participant in a patent application co-owned by Leiden University Medical Center (LUMC) and the University of Zurich on methods for training and using a machine learning model to predict the likelihood of an event in a patient with a malignant tumour by computational pathology (Application No. 23315438.4)

All other authors declare no competing interests.

## Author contributions

**Conception and design:** Simon Leedham, David N Church, Viktor H. Koelzer

**Financial support:** Joanne Edwards, Ian Tomlinson, Rachel Kerr, Simon Leedham, David N Church, Viktor H. Koelzer

**Administrative support:** Andrea Harkin, Leticia Campo, Viktor H. Koelzer

**Provision of study materials or patients:** Andrea Harkin, Karin Oien, Jennifer Hay, Joanne Edwards, Rachel Kerr, Viktor H. Koelzer

**Collection and assembly of data:** Konstantin Bräutigam, Henry Rotte, Enric Domingo, Nadia Nasreddin, Owen Sansom, Francesco Pezzella, Leticia Campo, Molly Browne, Mark Glaire, Andrea Harkin, Tim Iveson, Karin Oien, Noori Maka, Jennifer Hay, Joanne Edwards, Caroline Kelly, Rachel Kerr, David Kerr, David N Church, Viktor H. Koelzer

**Data analysis and interpretation:** Henry Rotte, Enric Domingo, Konstantin Bräutigam, Gabriel Valbuena, Tim Iveson, Mark Saunders, David Church, Viktor H. Koelzer

**Manuscript writing:** Konstantin Bräutigam, Henry Rotte, Enric Domingo, Viktor H. Koelzer

**Final approval of manuscript:** All authors

**Accountable for all aspects of the work:** All authors

## Acknowledgements

We thank the patients who participated in the SCOT trial and consented for their samples to be used for correlative research, as well as the recruiting clinicians and study team. We are also grateful to the Glasgow Tissue Research Facility (Glasgow University) for tissue microarray construction, Dr Esther Gil Vasquez for her contributions on the establishment of the dual color ISH protocol, and we thank the Translational Histopathology Laboratory (Department of Oncology, University of Oxford) for scanning.

## Declaration of generative AI and AI-assisted technologies

During the preparation of this work, the authors used DeepL Write (DeepL SE) and Claude Opus (Anthropic) to improve the language of the manuscript. Claude was additionally used to retrieve background information, to draft and revise specific sections. Generative AI was also used to assist in writing R code for statistical analyses and data visualization; all code was reviewed and validated by the authors, and all analyses were performed on the authors’ data. No generative AI was used to create, alter, or interpret primary data or images. The authors reviewed and extensively edited all AI-assisted content and take full responsibility for the content of the published article.

## Notes

### Clinical Trial

ISRCTN59757862

### Author Declarations

The SCOT (ISRCTN59757862) trial was conducted in accordance with the Declaration of Helsinki and Good Clinical Practice guidelines. The trial protocol received ethical approval by the West Glasgow Research Ethics Committee (REC reference 07/S0703/136; protocol version 1.1, January 21, 2008), with all subsequent amendments approved as required. All patients provided written informed consent before enrolment, including consent for the collection of tumour tissue and its use in translational research within the TransSCOT program. The present analysis was performed on tissue collected under this consent and falls within the original ethics approval.

