## Supplementary Appendix for "A regenerative stem cell phenotype marked by ANXA1 defines aggressive colorectal cancer biology"

### Supplementary Tables

#### Supplementary Tables S1A–D. Compartment analysis of ANXA1 / LGR5 expression

ANXA1 and LGR5 expression statistics by Consensus Molecular Subtype (imCMS), reported separately for the stroma and tumour compartments.

**Supplementary Table S1A.** *LGR5+ area in stroma compartment*

| Group | n | Median | IQR | Mean | SD |
|---|---|---|---|---|---|
| imCMS1 | 953 | 0.093 | 0.197 | 0.238 | 0.633 |
| imCMS2 | 1,418 | 0.080 | 0.171 | 0.539 | 3.048 |
| imCMS3 | 524 | 0.086 | 0.189 | 0.343 | 1.638 |
| imCMS4 | 892 | 0.075 | 0.152 | 0.228 | 0.865 |
| Mixed | 112 | 0.094 | 0.199 | 0.247 | 0.494 |

**Supplementary Table S1B.** *ANXA1+ area in stroma compartment*

| Group | n | Median | IQR | Mean | SD |
|---|---|---|---|---|---|
| imCMS1 | 953 | 0.993 | 1.612 | 1.639 | 2.205 |
| imCMS2 | 1,418 | 0.896 | 1.388 | 1.539 | 3.721 |
| imCMS3 | 524 | 0.969 | 1.377 | 1.476 | 1.697 |
| imCMS4 | 892 | 0.862 | 1.296 | 1.443 | 3.163 |
| Mixed | 112 | 0.958 | 1.313 | 1.554 | 1.927 |

**Supplementary Table S1C.** *LGR5+ area in tumour compartment*

| Group | n | Median | IQR | Mean | SD |
|---|---|---|---|---|---|
| imCMS1 | 953 | 0.435 | 2.174 | 2.232 | 5.004 |
| imCMS2 | 1,418 | 1.199 | 5.500 | 4.854 | 8.275 |
| imCMS3 | 524 | 0.776 | 3.778 | 3.647 | 6.621 |
| imCMS4 | 892 | 0.781 | 3.286 | 3.239 | 6.227 |
| Mixed | 112 | 0.871 | 3.182 | 3.318 | 5.294 |

**Supplementary Table S1D.** *ANXA1+ area in tumour compartment*

| Group | n | Median | IQR | Mean | SD |
|---|---|---|---|---|---|
| imCMS1 | 953 | 0.772 | 2.677 | 3.333 | 6.922 |
| imCMS2 | 1,418 | 0.300 | 0.717 | 1.156 | 3.986 |
| imCMS3 | 524 | 0.567 | 1.453 | 2.198 | 5.345 |
| imCMS4 | 892 | 0.372 | 1.214 | 1.849 | 5.554 |
| Mixed | 112 | 0.518 | 1.224 | 1.689 | 3.584 |

*n, sample size; IQR, interquartile range; SD, standard deviation. Values are reported to three decimal places. Median, mean, IQR, SD are noted as percentage (%) of respective compartment area.*

### Supplementary Table S2. ANXA1 expression in the invasive margin (IM) and clinical parameters.

*ANXA1, Annexin A1; CAPOX, capecitabine+oxaliplatin; CD, cluster of differentiation; FOLFOX, Folinic Acid+Fluorouracil (5-FU) +Oxaliplatin; IM, invasive margin; imCMS, image based consensus molecular subtype; LGR5, leucine-rich repeat-containing G-protein coupled receptor 5; MMR, mismatch repair; N, number (sample size); Q, quartile; RFI, recurrence-free interval; TC, tumour centre; Tumour proportion, equals 1-TSR*

| **Characteristic** | **Q1**<br>N = 472[a] | **Q2**<br>N = 472[a] | **Q3**<br>N = 472[a] | **Q4**<br>N = 472[a] | **p value**[b] |
|---|---|---|---|---|---|
| Age | 65 (58, 70) | 65 (58, 70) | 65 (59, 70) | 65 (58, 69) | 0.9 |
| **Sex** | | | | | 0.6 |
| Female | 193 (25%) | 188 (24%) | 188 (24%) | 205 (26%) | |
| Male | 279 (25%) | 284 (25%) | 284 (25%) | 267 (24%) | |
| Performance status | 127 (23%) | 164 (29%) | 127 (23%) | 138 (25%) | 0.025 |
| **Sidedness** | | | | | <0.001 |
| Left | 287 (26%) | 297 (27%) | 278 (25%) | 230 (21%) | |
| Right | 183 (24%) | 169 (22%) | 187 (24%) | 236 (30%) | |
| **Trial arm** | | | | | 0.7 |
| 12 weeks | 225 (24%) | 234 (25%) | 239 (25%) | 241 (26%) | |
| 24 weeks | 247 (26%) | 238 (25%) | 233 (25%) | 231 (24%) | |
| **Planned treatment** | | | | | 0.7 |
| CAPOX | 334 (25%) | 332 (25%) | 325 (24%) | 342 (26%) | |
| FOLFOX | 138 (25%) | 140 (25%) | 147 (26%) | 130 (23%) | |
| T4 stage | 142 (23%) | 148 (24%) | 139 (23%) | 180 (30%) | 0.016 |
| **Nodal stage** | | | | | 0.5 |
| 0 | 94 (25%) | 92 (24%) | 89 (23%) | 105 (28%) | |
| 1 | 276 (26%) | 261 (25%) | 259 (25%) | 248 (24%) | |
| 2 | 102 (22%) | 119 (26%) | 124 (27%) | 119 (26%) | |
| Tumour proportion | 45 (19, 75) | 15 (3, 38) | 7 (1, 25) | 6 (1, 25) | <0.001 |
| CD8, intraepithelial (z-score) | 0.16 (-0.37, 0.68) | 0.16 (-0.39, 0.72) | 0.22 (-0.38, 0.70) | 0.14 (-0.46, 0.63) | 0.6 |
| CD8, intrastromal (z-score) | 0.09 (-0.53, 0.68) | 0.12 (-0.48, 0.70) | 0.13 (-0.49, 0.69) | 0.13 (-0.44, 0.60) | 0.9 |
| FOXP3, intraepithelial (z-score) | 0.22 (-0.21, 0.57) | 0.21 (-0.25, 0.60) | 0.17 (-0.29, 0.58) | 0.17 (-0.36, 0.57) | 0.5 |
| FOXP3, intrastromal (z-score) | 0.21 (-0.46, 0.69) | 0.24 (-0.48, 0.76) | 0.27 (-0.48, 0.80) | 0.08 (-0.69, 0.59) | 0.004 |
| CD20, intraepithelial (z-score) | 0.21 (-0.47, 0.68) | 0.25 (-0.41, 0.74) | 0.22 (-0.58, 0.75) | 0.15 (-0.60, 0.70) | 0.4 |
| CD20, intrastromal (z-score) | 0.01 (-0.63, 0.54) | 0.23 (-0.43, 0.66) | 0.21 (-0.29, 0.69) | 0.12 (-0.37, 0.61) | <0.001 |
| CD68, intraepithelial (z-score) | -0.13 (-0.61, 0.50) | -0.05 (-0.62, 0.61) | 0.13 (-0.48, 0.76) | 0.18 (-0.48, 0.84) | <0.001 |
| CD68, intrastromal (z-score) | 0.09 (-0.38, 0.48) | 0.09 (-0.38, 0.50) | 0.15 (-0.36, 0.55) | 0.13 (-0.32, 0.51) | 0.4 |
| **FOXP3 (intrastromal) / CD8 (intraepithelial)** | | | | | 0.4 |
| high/high | 155 (26%) | 152 (25%) | 162 (27%) | 134 (22%) | |
| high/low | 170 (24%) | 175 (25%) | 178 (25%) | 178 (25%) | |
| low/low | 146 (25%) | 142 (25%) | 131 (23%) | 159 (28%) | |
| **imCMS** | | | | | <0.001 |
| imCMS1 | 103 (22%) | 100 (22%) | 119 (26%) | 143 (31%) | |
| imCMS2 | 223 (32%) | 170 (25%) | 172 (25%) | 128 (18%) | |
| imCMS3 | 53 (21%) | 78 (31%) | 52 (21%) | 69 (27%) | |
| imCMS4 | 87 (20%) | 108 (25%) | 111 (26%) | 120 (28%) | |
| Mixed | 6 (12%) | 16 (31%) | 18 (35%) | 12 (23%) | |
| **MMR status** | | | | | 0.035 |
| Deficient | 42 (21%) | 45 (22%) | 49 (24%) | 65 (32%) | |
| Proficient | 383 (27%) | 367 (26%) | 350 (24%) | 337 (23%) | |
| ***BRAF* V600E** | | | | | 0.4 |
| Mutant | 10 (22%) | 14 (30%) | 8 (17%) | 14 (30%) | |

| Characteristic | Q1<br>N = 472[a] | Q2<br>N = 472[a] | Q3<br>N = 472[a] | Q4<br>N = 472[a] | p value[b] |
|---|---|---|---|---|---|
| Wild-type | 88 (25%) | 84 (24%) | 95 (27%) | 84 (24%) | |
| ***KRAS*** | | | | | 0.2 |
| Mutant | 35 (26%) | 25 (19%) | 39 (29%) | 36 (27%) | |
| Wild-type | 63 (24%) | 73 (28%) | 64 (24%) | 62 (24%) | |

[a]Median (Q1, Q3); n (%). [b]Kruskal–Wallis rank sum test; Pearson's chi-squared test.

### Supplementary Table S3. LGR5 expression in the invasive margin (IM) and clinical parameters.

*ANXA1, Annexin A1; CAPOX, capecitabine+oxaliplatin; CD, cluster of differentiation; FOLFOX, Folinic Acid+Fluorouracil (5-FU) +Oxaliplatin; IM, invasive margin; imCMS, image based consensus molecular subtype; LGR5, leucine-rich repeat-containing G-protein coupled receptor 5; MMR, mismatch repair; N, number (sample size); Q, quartile; RFI, recurrence-free interval; TC, tumour centre; Tumour proportion, equals 1-TSR*

| **Characteristic** | **Q1**<br>N = 472[a] | **Q2**<br>N = 472[a] | **Q3**<br>N = 472[a] | **Q4**<br>N = 472[a] | **p value**[b] |
|---|---|---|---|---|---|
| Age | 64 (58, 70) | 65 (59, 71) | 64 (58, 69) | 65 (58, 70) | 0.6 |
| **Sex** | | | | | 0.9 |
| Female | 195 (25%) | 200 (26%) | 192 (25%) | 187 (24%) | |
| Male | 277 (25%) | 272 (24%) | 280 (25%) | 285 (26%) | |
| Performance status | 144 (26%) | 143 (26%) | 140 (25%) | 129 (23%) | 0.7 |
| **Sidedness** | | | | | 0.8 |
| Left | 271 (25%) | 267 (24%) | 276 (25%) | 278 (25%) | |
| Right | 197 (25%) | 202 (26%) | 189 (24%) | 187 (24%) | |
| **Trial arm** | | | | | >0.9 |
| 12 weeks | 240 (26%) | 234 (25%) | 231 (25%) | 234 (25%) | |
| 24 weeks | 232 (24%) | 238 (25%) | 241 (25%) | 238 (25%) | |
| **Planned treatment** | | | | | 0.4 |
| CAPOX | 336 (25%) | 346 (26%) | 329 (25%) | 322 (24%) | |
| FOLFOX | 136 (25%) | 126 (23%) | 143 (26%) | 150 (27%) | |
| T4 stage | 149 (24%) | 157 (26%) | 153 (25%) | 150 (25%) | >0.9 |
| **Nodal stage** | | | | | 0.8 |
| 0 | 98 (26%) | 91 (24%) | 97 (26%) | 94 (25%) | |
| 1 | 267 (26%) | 269 (26%) | 254 (24%) | 254 (24%) | |
| 2 | 107 (23%) | 112 (24%) | 121 (26%) | 124 (27%) | |
| Tumour proportion | 18 (2, 51) | 7 (1, 24) | 8 (1, 28) | 37 (14, 63) | <0.001 |
| CD8, intraepithelial (z-score) | 0.20 (-0.36, 0.69) | 0.17 (-0.34, 0.72) | 0.17 (-0.45, 0.73) | 0.11 (-0.45, 0.62) | 0.3 |
| CD8, intrastromal (z-score) | 0.14 (-0.50, 0.71) | 0.16 (-0.39, 0.76) | 0.09 (-0.46, 0.60) | 0.08 (-0.61, 0.57) | 0.018 |
| FOXP3, intraepithelial (z-score) | 0.12 (-0.31, 0.58) | 0.23 (-0.31, 0.60) | 0.20 (-0.24, 0.61) | 0.20 (-0.24, 0.54) | 0.8 |
| FOXP3, intrastromal (z-score) | 0.12 (-0.60, 0.68) | 0.28 (-0.45, 0.73) | 0.18 (-0.47, 0.72) | 0.20 (-0.50, 0.75) | 0.2 |
| CD20, intraepithelial (z-score) | 0.14 (-0.86, 0.65) | 0.20 (-0.52, 0.78) | 0.21 (-0.55, 0.69) | 0.27 (-0.32, 0.72) | 0.049 |
| CD20, intrastromal (z-score) | 0.10 (-0.57, 0.62) | 0.33 (-0.18, 0.78) | 0.14 (-0.39, 0.56) | 0.03 (-0.51, 0.57) | <0.001 |
| CD68, intraepithelial (z-score) | -0.02 (-0.59, 0.67) | -0.06 (-0.68, 0.69) | 0.15 (-0.51, 0.77) | 0.04 (-0.47, 0.61) | 0.079 |
| CD68, intrastromal (z-score) | 0.05 (-0.36, 0.53) | 0.14 (-0.37, 0.51) | 0.23 (-0.29, 0.56) | 0.06 (-0.40, 0.47) | 0.015 |
| **FOXP3 (intrastromal) / CD8 (intraepithelial)** | | | | | 0.4 |
| high/high | 148 (25%) | 164 (27%) | 143 (24%) | 148 (25%) | |
| high/low | 171 (24%) | 168 (24%) | 193 (28%) | 169 (24%) | |
| low/low | 151 (26%) | 137 (24%) | 135 (23%) | 155 (27%) | |
| **imCMS** | | | | | 0.017 |
| imCMS1 | 121 (26%) | 134 (29%) | 110 (24%) | 100 (22%) | |
| imCMS2 | 172 (25%) | 143 (21%) | 174 (25%) | 204 (29%) | |
| imCMS3 | 54 (21%) | 69 (27%) | 66 (26%) | 63 (25%) | |
| imCMS4 | 114 (27%) | 114 (27%) | 103 (24%) | 95 (22%) | |
| Mixed | 11 (21%) | 12 (23%) | 19 (37%) | 10 (19%) | |
| **MMR status** | | | | | 0.002 |
| Deficient | 55 (27%) | 62 (31%) | 53 (26%) | 31 (15%) | |
| Proficient | 350 (24%) | 338 (24%) | 356 (25%) | 393 (27%) | |
| ***BRAF* V600E** | | | | | 0.8 |
| Mutant | 15 (33%) | 9 (20%) | 10 (22%) | 12 (26%) | |

| Characteristic | Q1<br>N = 472[a] | Q2<br>N = 472[a] | Q3<br>N = 472[a] | Q4<br>N = 472[a] | p value[b] |
|---|---|---|---|---|---|
| Wild-type | 93 (26%) | 74 (21%) | 94 (27%) | 90 (26%) | |
| ***KRAS*** | | | | | >0.9 |
| Mutant | 36 (27%) | 30 (22%) | 33 (24%) | 36 (27%) | |
| Wild-type | 72 (27%) | 53 (20%) | 71 (27%) | 66 (25%) | |

[a]Median (Q1, Q3); n (%). [b]Kruskal–Wallis rank sum test; Pearson’s chi-squared test.

### Supplementary Table S4. ANXA1-LGR5 quartiles in the invasive margin (IM) and clinical data.

*ANXA1, Annexin A1; CAPOX, capecitabine+oxaliplatin; CD, cluster of differentiation; FOLFOX, Folinic Acid+Fluorouracil (5-FU) +Oxaliplatin; IM, invasive margin; imCMS, image based consensus molecular subtype; LGR5, leucine-rich repeat-containing G-protein coupled receptor 5; MMR, mismatch repair; N, number (sample size); Q, quartile; RFI, recurrence-free interval; TC, tumour centre; Tumour proportion, equals 1-TSR*

| **Characteristic** | **Q1**<br>N = 472[a] | **Q2**<br>N = 472[a] | **Q3**<br>N = 472[a] | **Q4**<br>N = 472[a] | **p value**[b] |
|---|---|---|---|---|---|
| Age | 65 (58, 70) | 64 (58, 69) | 64 (58, 70) | 65 (59, 70) | 0.6 |
| **Sex** | | | | | 0.6 |
| Female | 187 (24%) | 202 (26%) | 186 (24%) | 199 (26%) | |
| Male | 285 (26%) | 270 (24%) | 286 (26%) | 273 (25%) | |
| Performance status | 127 (23%) | 147 (26%) | 139 (25%) | 143 (26%) | 0.5 |
| **Sidedness** | | | | | <0.001 |
| Left | 290 (27%) | 294 (27%) | 280 (26%) | 228 (21%) | |
| Right | 176 (23%) | 176 (23%) | 186 (24%) | 237 (31%) | |
| **Trial arm** | | | | | 0.9 |
| 12 weeks | 233 (25%) | 228 (24%) | 241 (26%) | 237 (25%) | |
| 24 weeks | 239 (25%) | 244 (26%) | 231 (24%) | 235 (25%) | |
| **Planned treatment** | | | | | 0.6 |
| CAPOX | 339 (25%) | 333 (25%) | 322 (24%) | 339 (25%) | |
| FOLFOX | 133 (24%) | 139 (25%) | 150 (27%) | 133 (24%) | |
| T4 stage | 147 (24%) | 131 (22%) | 152 (25%) | 179 (29%) | 0.009 |
| **Nodal stage** | | | | | 0.6 |
| 0 | 88 (23%) | 93 (24%) | 94 (25%) | 105 (28%) | |
| 1 | 264 (25%) | 264 (25%) | 271 (26%) | 245 (23%) | |
| 2 | 120 (26%) | 115 (25%) | 107 (23%) | 122 (26%) | |
| Positive lymph nodes | 2 (1, 4) | 1 (1, 3) | 2 (1, 3) | 1 (1, 4) | 0.8 |
| RFI event | 94 (21%) | 114 (26%) | 107 (24%) | 129 (29%) | 0.057 |
| Liver metastasis | 28 (17%) | 51 (31%) | 40 (24%) | 48 (29%) | 0.040 |
| Lung metastasis | 28 (21%) | 31 (24%) | 36 (27%) | 36 (27%) | 0.7 |
| Bone metastasis | 1 (8.3%) | 3 (25%) | 4 (33%) | 4 (33%) | 0.6 |
| Peritoneal metastasis | 14 (25%) | 9 (16%) | 10 (18%) | 22 (40%) | 0.049 |
| Local nodal metastasis | 10 (20%) | 12 (24%) | 10 (20%) | 17 (35%) | 0.4 |
| Regional nodal metastasis | 8 (16%) | 17 (33%) | 11 (22%) | 15 (29%) | 0.3 |
| Distant nodal metastasis | 8 (25%) | 11 (34%) | 4 (13%) | 9 (28%) | 0.3 |
| Other metastasis | 23 (23%) | 24 (24%) | 25 (25%) | 28 (28%) | 0.9 |
| Tumour proportion | 51 (27, 75) | 17 (4, 42) | 5 (1, 18) | 4 (1, 20) | <0.001 |
| CD8, intraepithelial (z-score) | 0.18 (-0.34, 0.65) | 0.10 (-0.47, 0.72) | 0.30 (-0.34, 0.74) | 0.12 (-0.47, 0.61) | 0.074 |
| CD8, intrastromal (z-score) | 0.10 (-0.58, 0.67) | 0.07 (-0.53, 0.70) | 0.15 (-0.41, 0.70) | 0.14 (-0.44, 0.62) | 0.6 |
| FOXP3, intraepithelial (z-score) | 0.23 (-0.18, 0.56) | 0.22 (-0.25, 0.62) | 0.17 (-0.27, 0.58) | 0.17 (-0.39, 0.57) | 0.2 |
| FOXP3, intrastromal (z-score) | 0.24 (-0.45, 0.75) | 0.17 (-0.48, 0.70) | 0.25 (-0.45, 0.77) | 0.09 (-0.70, 0.60) | 0.006 |
| CD20, intraepithelial (z-score) | 0.23 (-0.34, 0.68) | 0.26 (-0.50, 0.73) | 0.21 (-0.77, 0.77) | 0.13 (-0.60, 0.71) | 0.4 |
| CD20, intrastromal (z-score) | -0.01 (-0.65, 0.47) | 0.23 (-0.41, 0.66) | 0.26 (-0.25, 0.74) | 0.15 (-0.40, 0.63) | <0.001 |
| CD68, intraepithelial (z-score) | -0.05 (-0.49, 0.51) | -0.06 (-0.62, 0.64) | 0.05 (-0.60, 0.70) | 0.17 (-0.53, 0.89) | 0.016 |
| CD68, intrastromal (z-score) | 0.09 (-0.39, 0.49) | 0.09 (-0.37, 0.51) | 0.13 (-0.34, 0.54) | 0.12 (-0.35, 0.52) | 0.5 |
| **FOXP3 (intrastromal) / CD8 (intraepithelial)** | | | | | 0.038 |
| high/high | 166 (28%) | 137 (23%) | 169 (28%) | 131 (22%) | |
| high/low | 164 (23%) | 181 (26%) | 177 (25%) | 179 (26%) | |
| low/low | 142 (25%) | 150 (26%) | 125 (22%) | 161 (28%) | |
| **imCMS** | | | | | <0.001 |
| imCMS1 | 92 (20%) | 107 (23%) | 119 (26%) | 147 (32%) | |

| **Characteristic** | **Q1**<br>N = 472[a] | **Q2**<br>N = 472[a] | **Q3**<br>N = 472[a] | **Q4**<br>N = 472[a] | **p value**[b] |
|---|---|---|---|---|---|
| imCMS2 | 224 (32%) | 184 (27%) | 162 (23%) | 123 (18%) | |
| imCMS3 | 59 (23%) | 66 (26%) | 65 (26%) | 62 (25%) | |
| imCMS4 | 90 (21%) | 102 (24%) | 107 (25%) | 127 (30%) | |
| Mixed | 7 (13%) | 13 (25%) | 19 (37%) | 13 (25%) | |
| **MMR status** | | | | | 0.001 |
| Deficient | 35 (17%) | 52 (26%) | 45 (22%) | 69 (34%) | |
| Proficient | 397 (28%) | 361 (25%) | 346 (24%) | 333 (23%) | |
| ***BRAF* V600E** | | | | | 0.3 |
| Mutant | 13 (28%) | 12 (26%) | 7 (15%) | 14 (30%) | |
| Wild-type | 77 (22%) | 92 (26%) | 95 (27%) | 87 (25%) | |
| ***KRAS*** | | | | | 0.7 |
| Mutant | 29 (21%) | 32 (24%) | 35 (26%) | 39 (29%) | |
| Wild-type | 61 (23%) | 72 (27%) | 67 (26%) | 62 (24%) | |

[a]Median (Q1, Q3); n (%).

[b]Kruskal–Wallis rank sum test; Pearson's chi-squared test; Fisher's exact test.

### Table S5: Multivariable analysis for invasive margin (IM) samples.

*FOLFOX*: Folinic Acid+Fluorouracil (5-FU)+Oxaliplatin; *M*: Male; *MMR*: mismatch repair; *N*: nodal; *Q*: quartile

| | OS (N=1620) | | CSS (N=1569) | | RFI (N=1620) | | SAR (N=380) | |
|---|---|---|---|---|---|---|---|---|
| **Variable** | **HR (CIs)** | **p** | **HR (CIs)** | **p** | **HR (CIs)** | **p** | **HR (CIs)** | **p** |
| ANXA1-LGR5 Q1 vs Q2 | 1.16 (0.83-1.63) | 0.387 | 1.51 (0.98-2.34) | 0.062 | 1.35 (0.99-1.84) | 0.058 | 1.05 (0.71-1.53) | 0.814 |
| ANXA1-LGR5 Q1 vs Q3 | 1.09 (0.76-1.56) | 0.634 | 1.51 (0.97-2.37) | 0.070 | 1.22 (0.87-1.70) | 0.244 | 0.91 (0.61-1.36) | 0.643 |
| ANXA1-LGR5 Q1 vs Q4 | 1.28 (0.90-1.82) | 0.172 | 1.55 (0.99-2.44) | 0.055 | 1.35 (0.97-1.88) | 0.075 | 0.99 (0.68-1.45) | 0.968 |
| Tumour proportion* | 1.00 (0.99-1.00) | 0.436 | 1.00 (0.99-1.00) | 0.435 | 1.00 (0.99-1.00) | 0.317 | 1.00 (0.99-1.00) | 0.436 |
| T4 stage | 2.06 (1.64-2.58) | <0.001 | 2.65 (2.01-3.49) | <0.001 | 2.06 (1.67-2.54) | <0.001 | 1.48 (1.15-1.89) | 0.002 |
| N stage | 1.67 (1.42-1.98) | <0.001 | 2.07 (1.68-2.54) | <0.001 | 1.78 (1.53-2.08) | <0.001 | 1.28 (1.06-1.56) | 0.012 |
| Sidedness Right | 1.47 (1.16-1.86) | 0.001 | 1.73 (1.29-2.31) | <0.001 | 1.40 (1.13-1.74) | 0.002 | 1.51 (1.16-1.96) | 0.002 |
| Arm 24 weeks | 0.93 (0.75-1.16) | 0.542 | 0.85 (0.65-1.12) | 0.244 | 0.90 (0.73-1.10) | 0.307 | 1.16 (0.90-1.49) | 0.255 |
| Planned treatment Folfox | 0.97 (0.76-1.24) | 0.811 | 0.95 (0.71-1.28) | 0.738 | 1.05 (0.85-1.31) | 0.638 | 1.00 (0.76-1.30) | 0.986 |
| Sex M | 0.99 (0.78-1.24) | 0.897 | 0.75 (0.57-0.99) | 0.043 | 0.95 (0.77-1.18) | 0.660 | 0.75 (0.59-0.97) | 0.027 |
| Age at registration | 1.02 (1.00-1.03) | 0.010 | 1.01 (0.99-1.02) | 0.360 | 1.00 (0.98-1.01) | 0.445 | 1.02 (1.01-1.04) | 0.002 |
| MMR IHC loss | 0.67 (0.46-0.98) | 0.040 | 0.60 (0.38-0.95) | 0.030 | 0.56 (0.38-0.82) | 0.003 | 1.33 (0.84-2.09) | 0.223 |

**Tumour proportion as a continuous variable* is printed in italics: *Tumour proportion as a continuous variable

### Supplementary Figures

#### Supplementary Figure S1

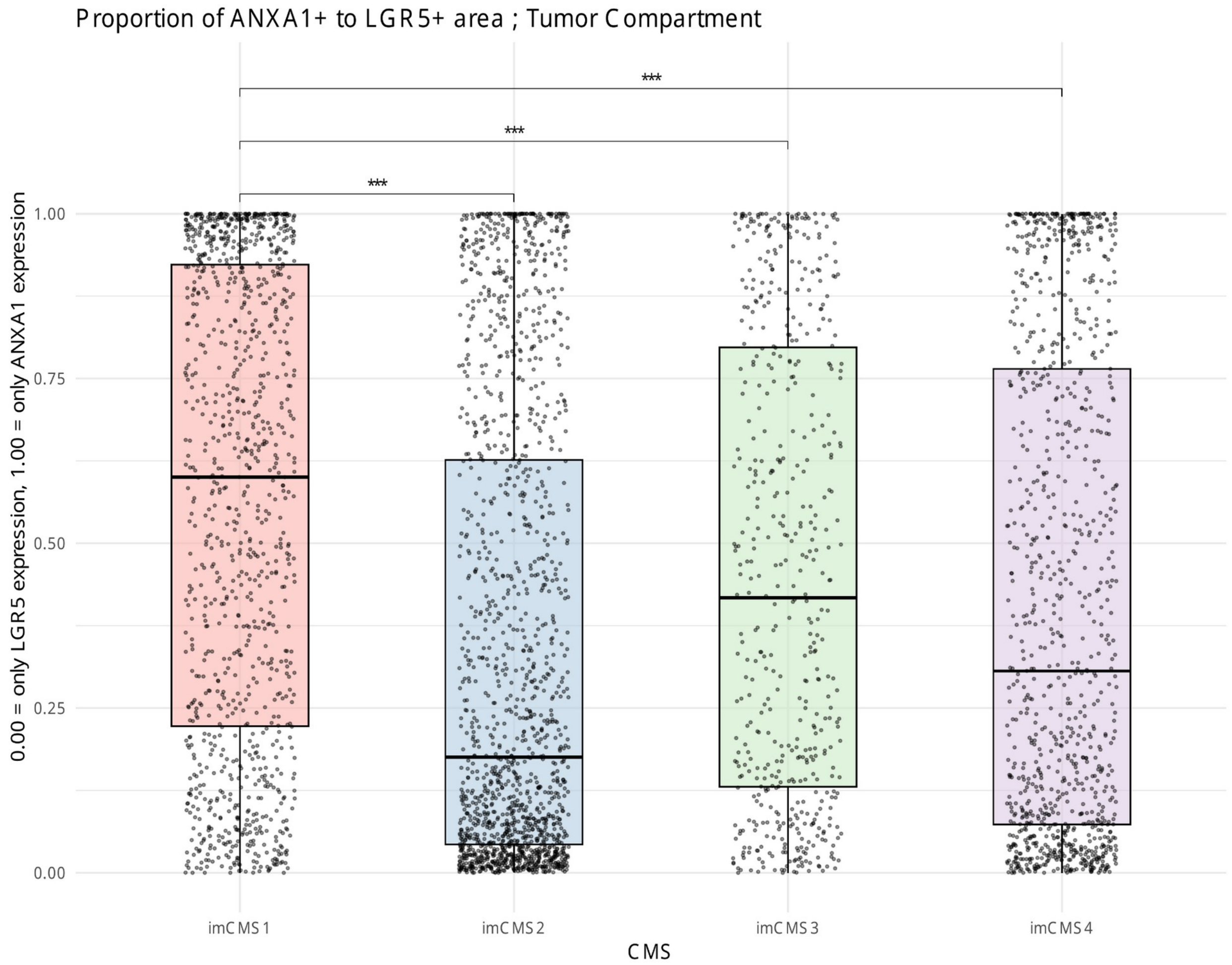


S1: proportion of ANXA1+ area out of total ANXA1+ plus LGR5+ area in the tumour compartment, by imCMS (0 = LGR5 only, 1 = ANXA1 only).

**Supplementary Figure S2A**

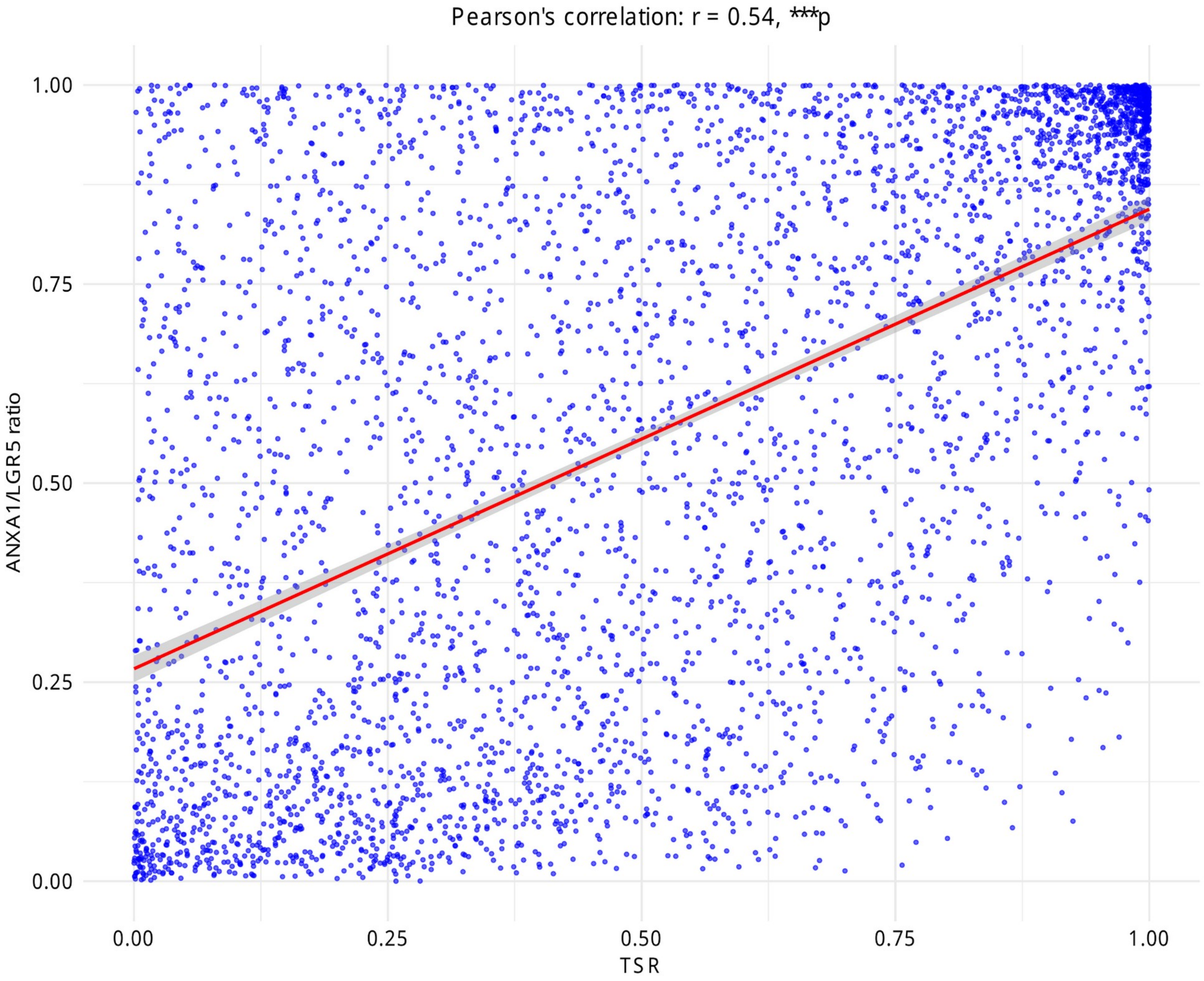


S2A: ANXA1/LGR5 proportion against tumour–stroma ratio (TSR), with Pearson's r.

**Supplementary Figure S2B**

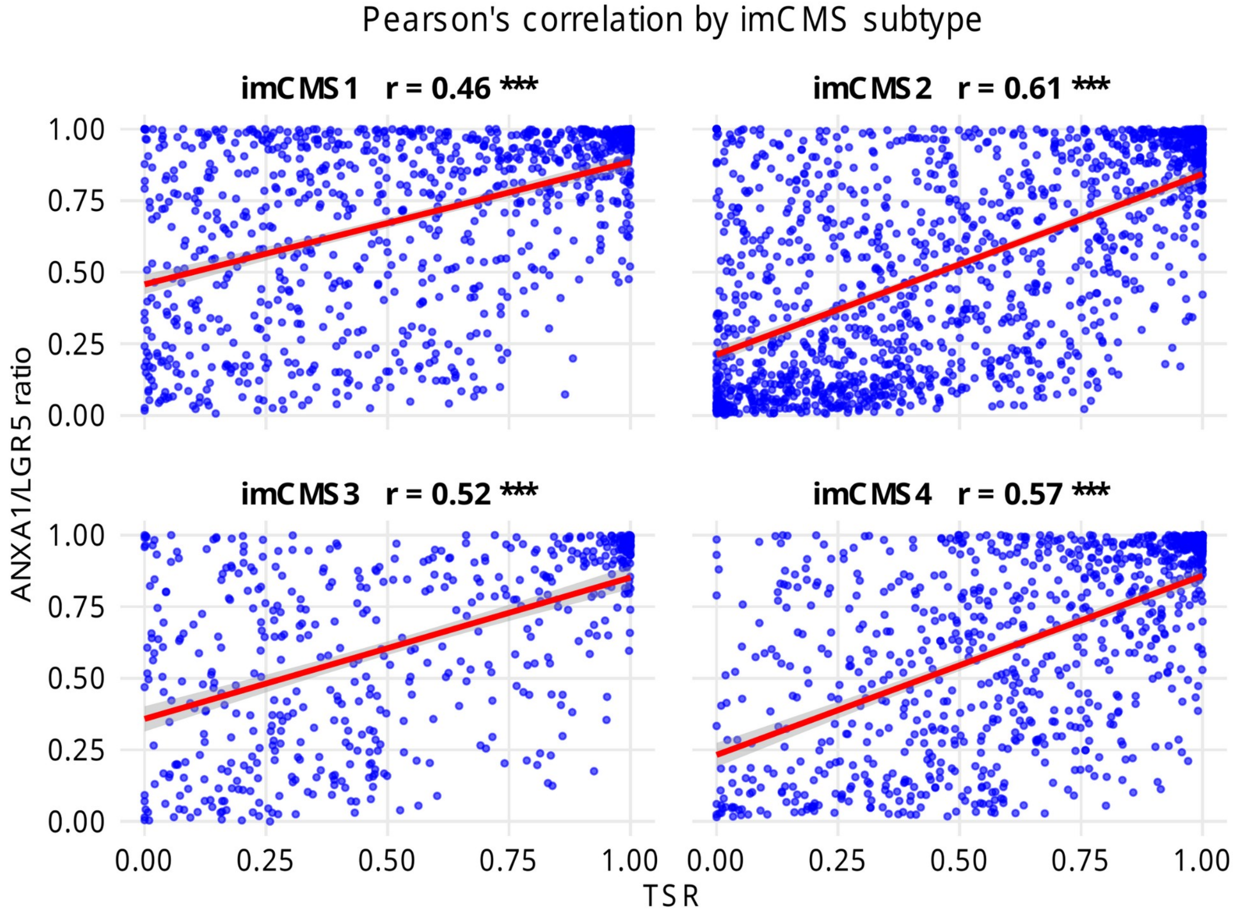


S2B: ANXA1/LGR5 proportion against tumour–stroma ratio (TSR), with Pearson’s r, split by imCMS subtype.

**Supplementary Figure S3A**

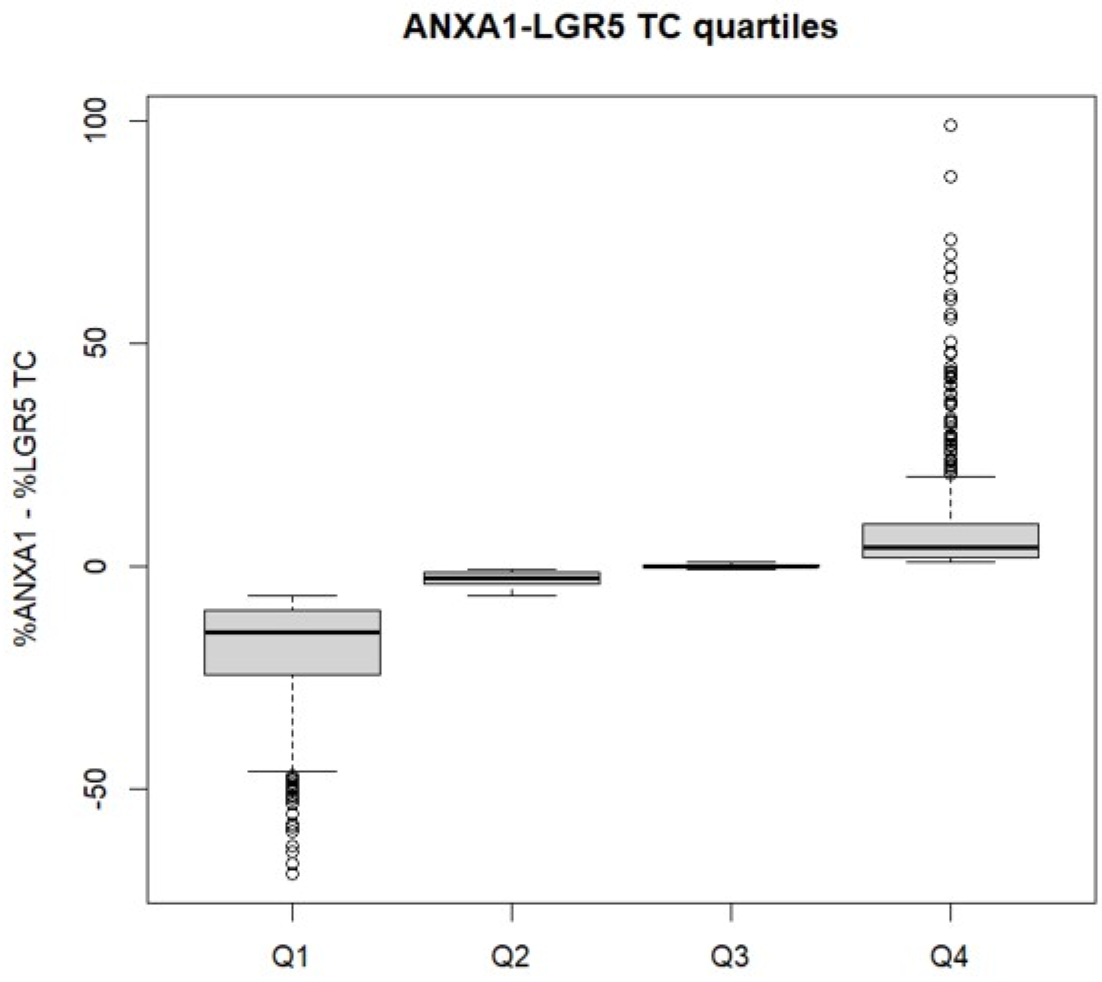


S3A: distribution of ANXA1 − LGR5 (%) across quartiles, tumour centre (TC).

**Supplementary Figure S3B**

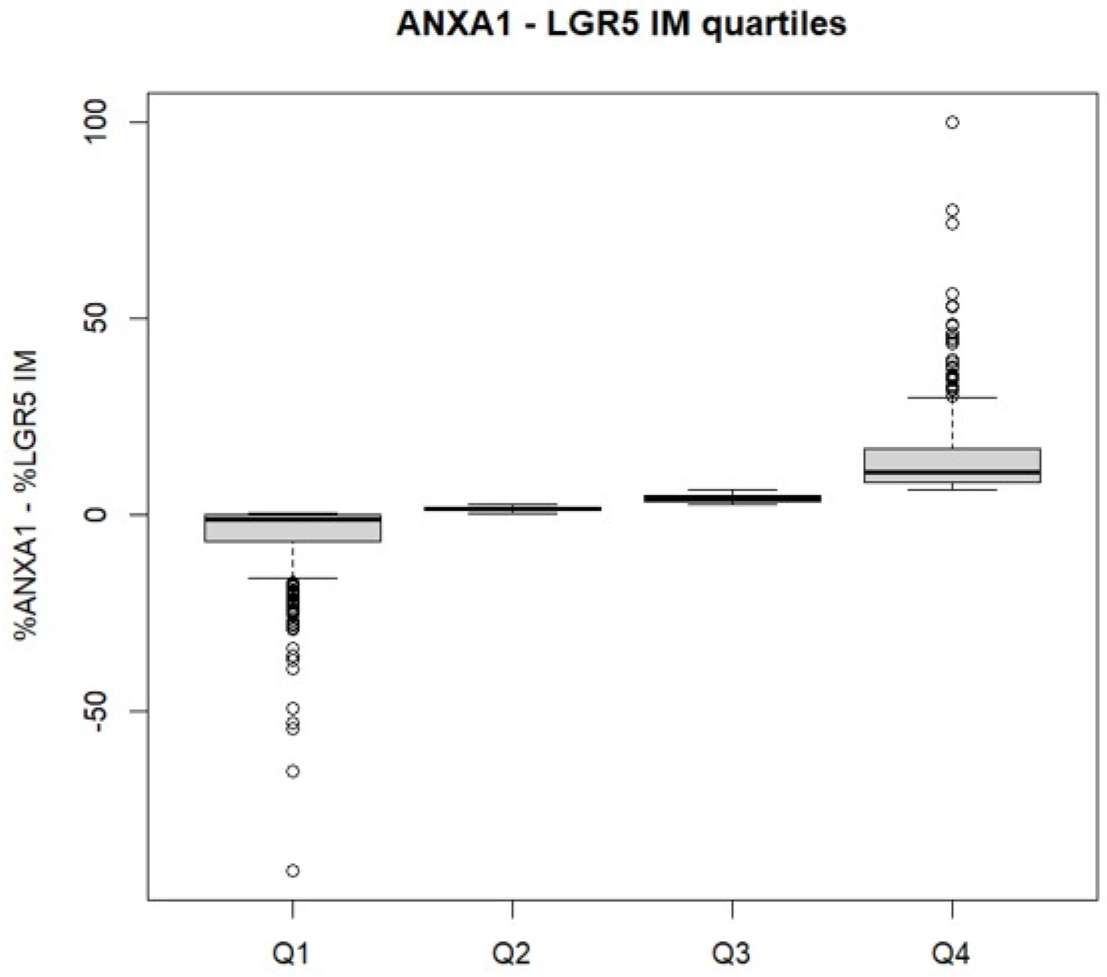


S3B: distribution of ANXA1 − LGR5 (%) across quartiles, invasive margin (IM).

**Supplementary Figure S4A**

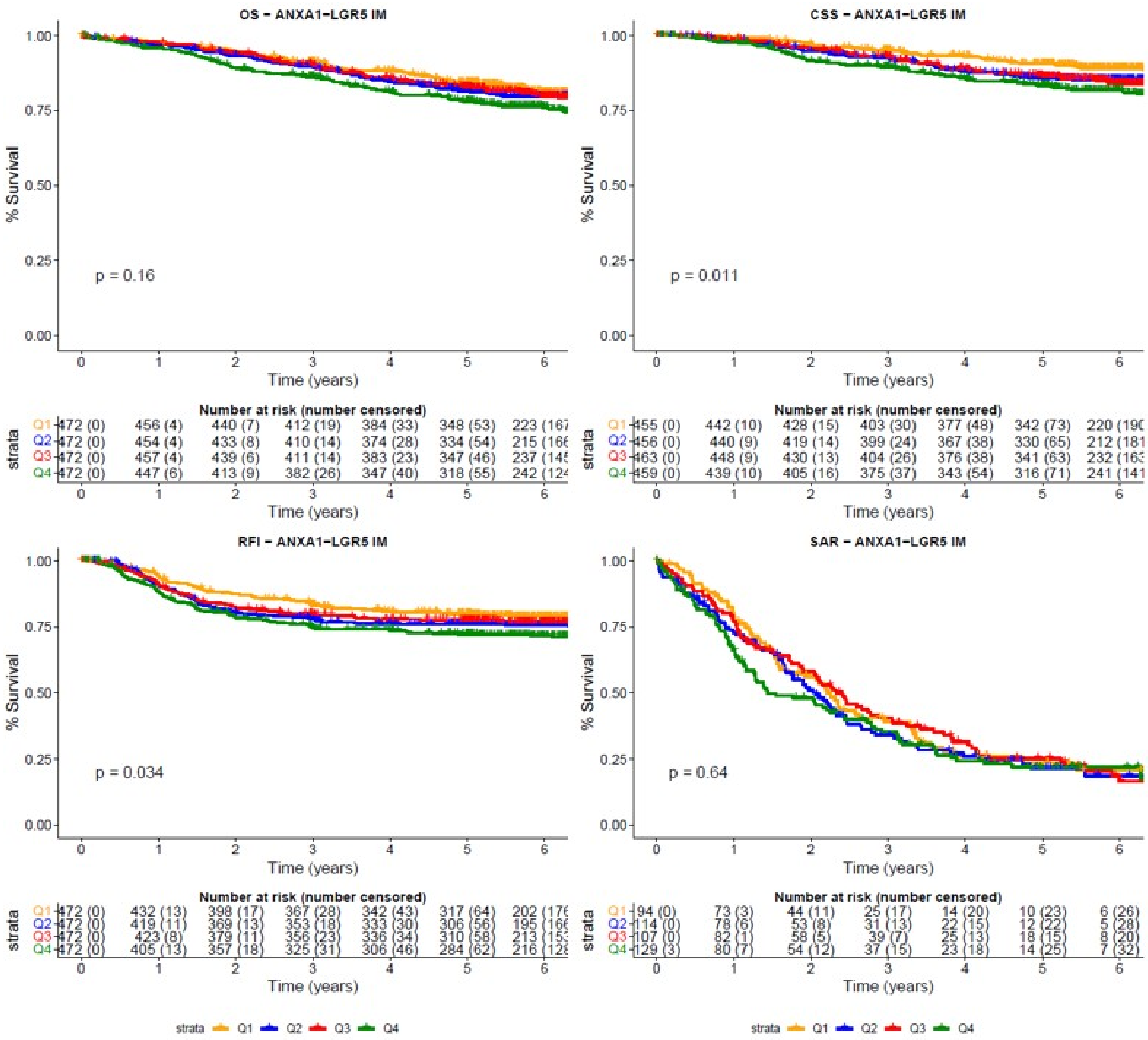


S4A: Kaplan–Meier curves for OS, CSS, RFI and SAR in IM samples, by ANXA1−LGR5 quartile

**Supplementary Figure S4B**

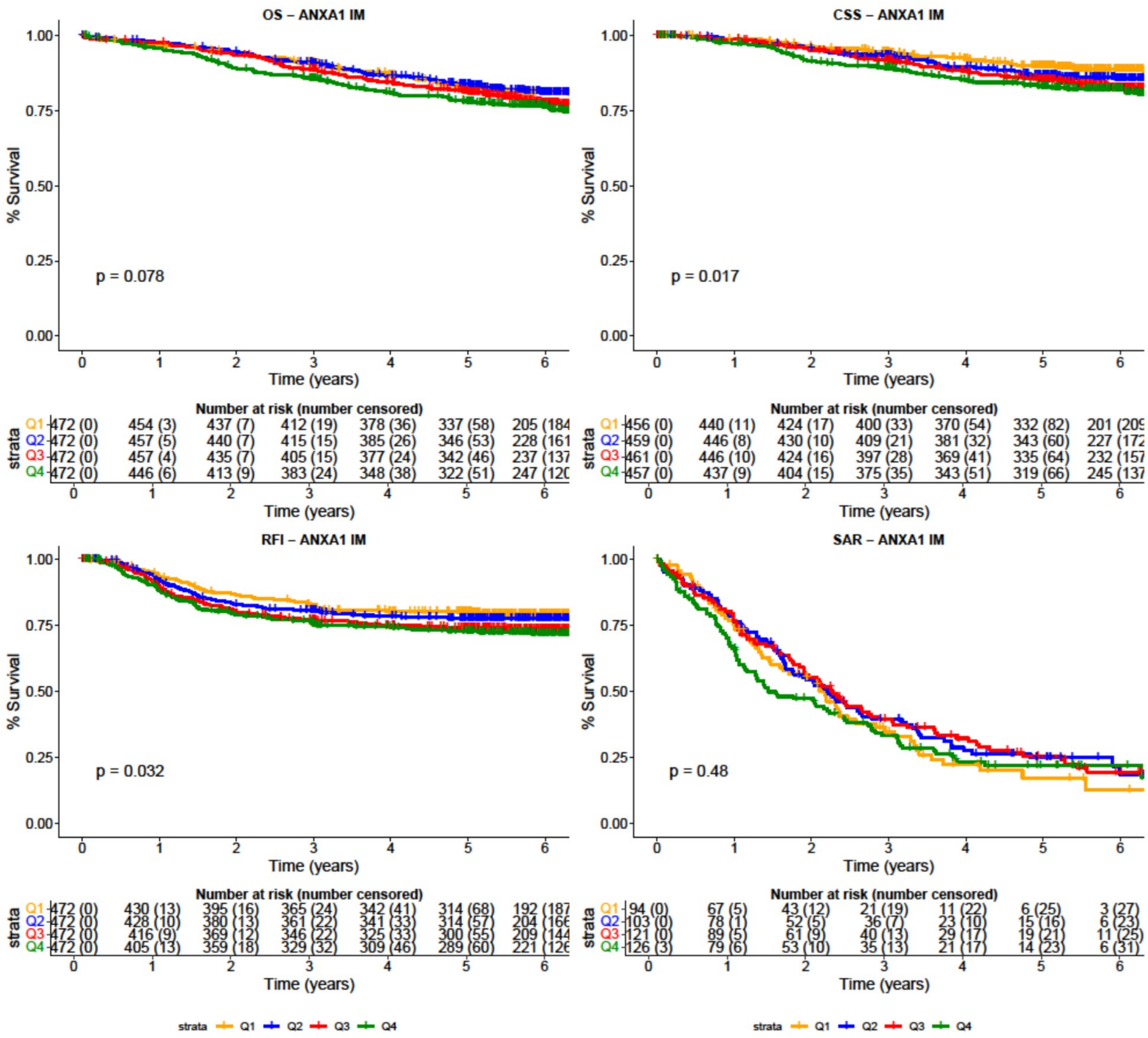


S4B: Kaplan–Meier curves for OS, CSS, RFI and SAR in IM samples, by ANXA1 quartile

**Supplementary Figure S4C**

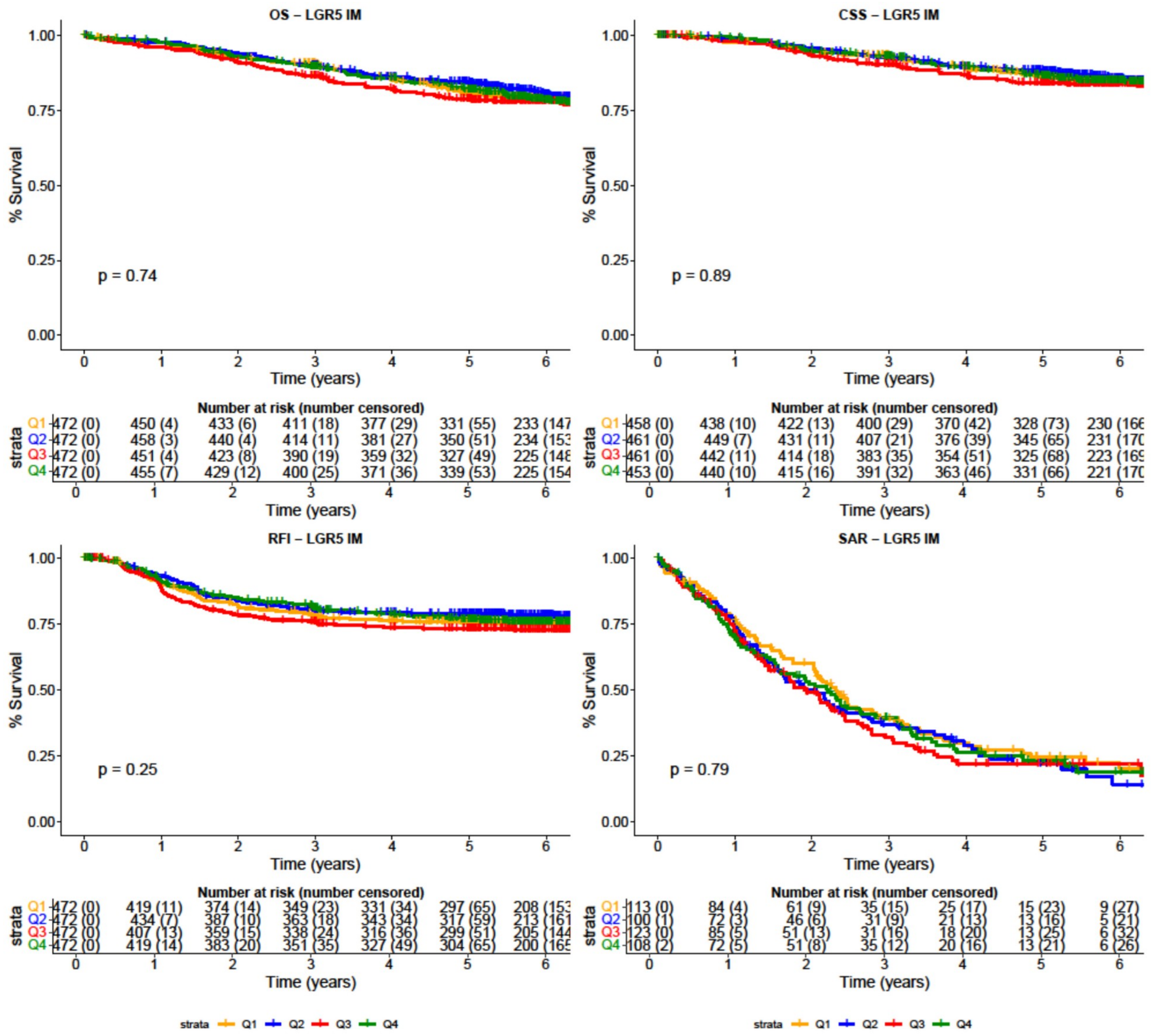


S4C: Kaplan–Meier curves for OS, CSS, RFI and SAR in IM samples, by LGR5 quartile

### Supplementary Figure S5A

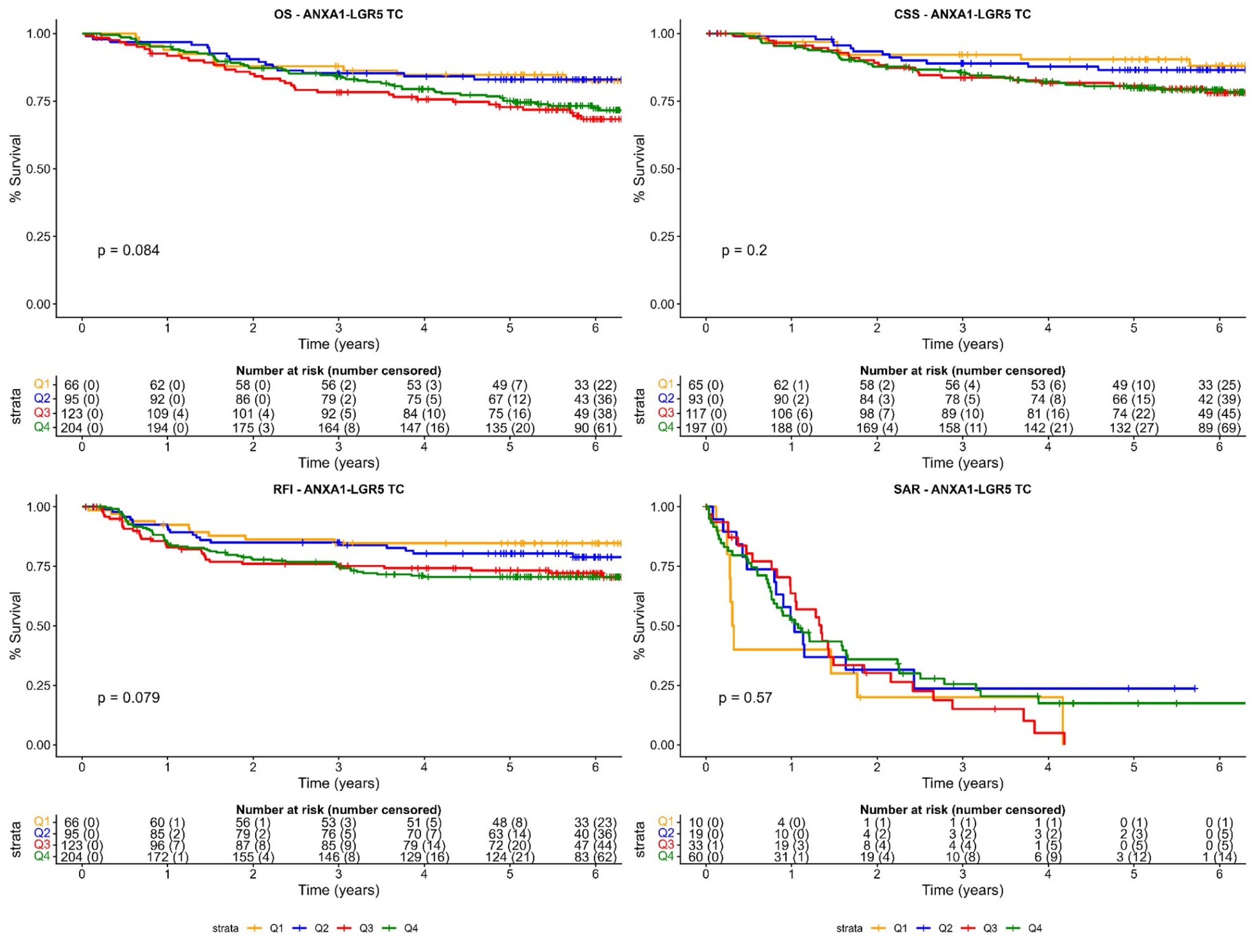


S5A: Kaplan–Meier curves by ANXA1−LGR5 quartile in TC samples, restricted to imCMS1.

### Supplementary Figure S5B

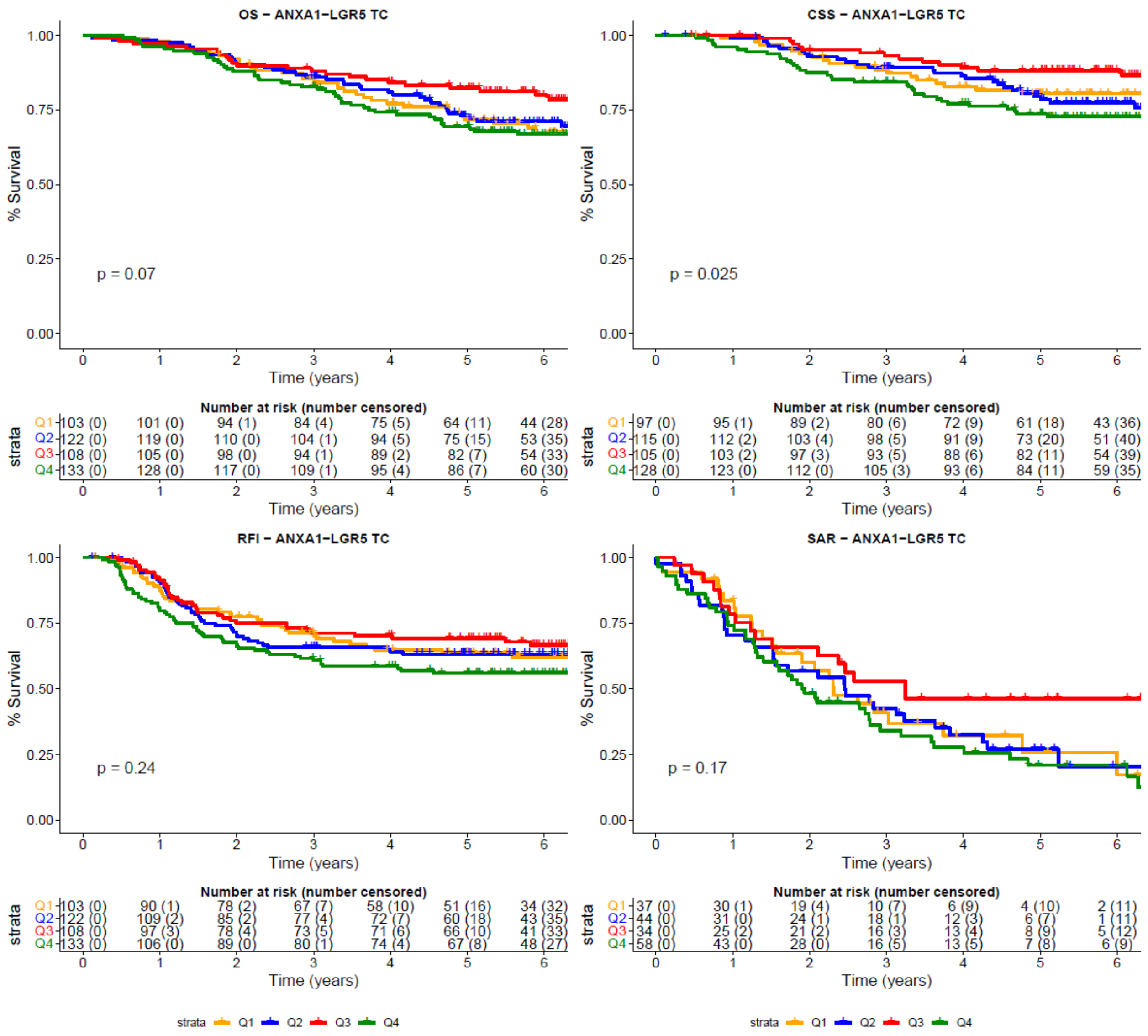


S5B: Kaplan–Meier curves by ANXA1−LGR5 quartile in TC samples, restricted to imCMS4.
